# Modelling the adjustment of COVID-19 response and exit from dynamic zero-COVID in China

**DOI:** 10.1101/2022.12.14.22283460

**Authors:** Kathy Leung, Gabriel M. Leung, Joseph T. Wu

**Author notes:** Correspondence: Kathy Leung. Contributed equally.

## Abstract

**Background:** Since the initial Wuhan outbreak, China has been containing COVID-19 outbreaks through its “dynamic zero-COVID” policy. Striking a balance between sustainability and cost-benefit, China has recently begun to adjust its COVID-19 response strategies, e.g. by announcing the “20 measures” on 11 November and further the “10 measures” on 7 December 2022. Strategies for safely exiting from dynamic zero-COVID (i.e. without catastrophically overburdening health systems and/or incurring unacceptably excessive morbidity and mortality) are urgently needed.

**Methods:** We use simulations to assess the respective and combined effectiveness of fourth-dose heterologous boosting, large-scale antiviral treatment and public health and social measures (PHSMs) that might allow China to further adjust COVID-19 response and exit from zero-COVID safely after 7 December 2022. We also assess whether local health systems can cope with the surge of COVID-19 cases posed by reopening, given that *chunyun*, a 40-day period with extremely high mobility across China associated with Spring Festival, will begin on 7 January 2023.

**Findings:** Reopening against Omicron transmission should be supported by the following interventions: 1) fourth-dose heterologous boosting 30-60 days before reopening by vaccinating 4-8% of the population per week with ≥85% uptake across all ages; 2) timely antiviral treatment with ≥60% coverage; 3) moderate PHSMs to reduce transmissibility by 47-69%. With fourth-dose vaccination coverage of 85% and antiviral coverage of 60%, the cumulative mortality burden would be reduced by 26-35% to 448-503 per million, compared with reopening without any of these interventions. Simultaneously reopening all provinces under current PHSMs would still lead to hospitalisation demand that are 1.5-2.5 times of surge hospital capacity (2.2 per 10,000 population per day).

**Interpretation:** Although the surge of disease burden posed by reopening in December 2022 – January 2023 would likely overload many local health systems across the country, the combined effect of vaccination, antiviral treatment and PHSMs could substantially reduce COVID-19 morbidity and mortality as China transits from dynamic-zero to normality. Planning for such a nationwide, coordinated reopening should be an urgent priority as part of the global exit from the acute phase of the COVID-19 pandemic.

**Funding:** COVID-19 Vaccines Evaluation Program, Chinese Center for Disease Control and Prevention; Health and Medical Research Fund, Health Bureau, The Government of the Hong Kong SAR; General Research Fund, Research Grants Council, Hong Kong

**Research in context:** *Evidence before this study:* We searched PubMed and preprint archives for articles published up to 7 December 2021, that contained information about exit strategies of zero-COVID or reopening in China after the emergence of Omicron using the terms “China”, “Omicron”, “B.1.1.529”, “COVID-19”, “SARS-CoV-2”, “vaccin*”, “vaccine”, “antiviral”, “control measures”, “non-pharmaceutical intervention”, “public health and social measure”, “zero-COVID”, “exit strategy” and “reopen*”. We only found one study by Wang et al (doi: 10.1101/2022.05.07.22274792) but they assessed the feasibility of sustaining SARS-CoV-2 containment with zero-COVID strategy in China. To our knowledge, there is no discussion of exit strategies of the zero-COVID strategy or assessment of feasibility of reopening in China.

*Added value of this study:* Reopening against Omicron transmission should be supported by the following interventions: 1) fourth-dose heterologous boosting 30-60 days before reopening by vaccinating 4-8% of the population per week with ≥85% uptake across all ages; 2) timely antiviral treatment with ≥60% coverage; 3) moderate PHSMs to reduce transmissibility by 47-69%. With fourth-dose vaccination coverage of 85% and antiviral coverage of 60%, the cumulative mortality burden would be reduced by 26-35% to 448-503 per million, compared with reopening without any of these interventions. Simultaneously reopening all provinces under current PHSMs would still lead to hospitalisation demand that are 1.5-2.5 times of surge hospital capacity (2.2 per 10,000 population per day).

*Implications of all the available evidence:* Although the surge of disease burden posed by reopening in December 2022 – January 2023 would likely overload many local health systems across the country, the combined effect of vaccination, antiviral treatment and PHSMs could substantially reduce COVID-19 morbidity and mortality as China transits from dynamic-zero to normality. Planning for such a nationwide, coordinated reopening should be an urgent priority as part of the global exit from the acute phase of the COVID-19 pandemic.

## Introduction

Since March 2020, China has been containing successive sporadic clusters of COVID-19 infection through its “dynamic zero-COVID” policy – by mass lockdowns, universal compulsory testing of entire districts or even whole cities, stringent quarantine and isolation enforced down to the neighbourhood level, universal digital contact tracing and importation controls ^1,2^. Although this zero-COVID strategy had remained effective in 2020-2021, the emergence and spread of Omicron since early 2022 has triggered prolonged lockdowns and major disruptions in megacities such as Beijing and Shanghai, with ramifications far beyond its own shores nationally and globally in terms of the economy, trade, and commerce. Striking a balance between sustainability and cost-benefit of dynamic-zero, China has recently begun to adjust its COVID-19 response strategies, notably by announcing the “20 measures” on 11 November and further the “10 measures” on 7 December 2022 ^3,4^.

Elsewhere in other parts of the world, the majority of countries are transiting to a “living with the virus” strategy, having built up considerable population immunity through multiple infection waves of ancestral strain and variants of concern (VOCs) and with the majority population having received two or three (sometimes even four) doses of vaccines. Given the interdependency of infectious disease dynamics at the meta-population level, when and how China can safely exit from its dynamic zero-COVID policy (in terms of significant morbidity and mortality as well as exceedance of the surge capacity of local health systems) has important implications to health security and economic stability globally.

As of 6 December 2022, mainland China has tallied just over 349,938 confirmed COVID-19 cases and 5,235 COVID-related deaths. Before the mass vaccination programme, the seroprevalence was <5% in Wuhan and <1% outside Wuhan in Hubei, even though they were the epicentres of the first wave in 2020 ^5^. As such, existing population immunity nationwide is primarily conferred by passive immunisation with the domestically produced inactivated virus vaccines. As of 28 November 2022, the vaccine uptake of two- and three-dose vaccination are 91% and 57%, respectively. Although three-dose homologous vaccinations are highly effective in reducing the risk of Omicron hospitalisation and death, they have limited lasting impact in boosting immunity against Omicron (or future VOCs) transmission. Recent data suggests that for individuals who have received two doses of inactivated vaccines, a third dose of a heterologous vaccine increases neutralising antibodies to levels that correlate with substantial reduction in susceptibility to Omicron infection ^6^. Thus, our basic premise is that high uptake of three-dose inactivated vaccines followed by heterologous boosters (i.e., fourth dose) would provide significant protection against Omicron transmission for 2-3 months to create a time window for safer reopening.

Timely antiviral treatment of symptomatic Omicron patients has now been proven to be effective in substantially reducing their risk of hospitalisation and death ^7^. Nirmatrelvir/Ritonavir has already been approved for treating COVID in mainland China whilst five domestic companies are manufacturing Nirmatrelvir/Ritonavir for exports ^8^. In addition, several domestic candidate oral drugs, including SIM0417 (a 3CL protease inhibitor similar to Nirmatrelvir), are undergoing Phase 2/3 trials. In this study, we assume that large-scale antiviral treatments are widely available by the time of reopening.

Here, we investigate to what extent optimal deployment of vaccines, antivirals and public health and social measures (PHSMs) could allow China to further adjust COVID-19 response and exit from zero-COVID safely after 7 December 2022. We also assess whether the local health systems can cope with the surge of disease burden posed by reopening, given that *chunyun*, a 40-day period with extremely high mobility across China associated with Spring Festival, will begin on 7 January 2023.

## Methods

### Vaccine effectiveness (VE) of 4^th^-dose heterologous boosting

As of 28 November 2022, the two- and three-dose vaccine uptake in mainland China was 91% and 57%. Since more than 95% of vaccines administered are inactivated virus vaccines, we assume that all vaccinees would receive inactivated virus vaccines for their first three doses. Following the widely used method developed by Khoury et al for predicting vaccine effectiveness from neutralisation antibody titres, we estimate that several domestically-made vaccines have substantial VE in reducing Omicron transmission (for a limited duration) if given as 4^th^-dose heterologous booster (see **Table S1** and **Appendix** for details) ^9^. We assume that VEs conferred by 4^th^-dose boosting in reducing hospitalisations and deaths are equivalent to that from three doses of inactivated virus vaccines against the Omicron wave in Hong Kong (**Table S2**) ^10^.

We use a “leaky” vaccine action model to estimate vaccine-induced population immunity in reducing susceptibility to infection, infectiousness, hospitalisation, and death as a function of 4^th^-dose uptake over time (**Figure 1**). We assume that 4^th^-dose heterologous boosting would be delivered via two hypothetical vaccines A and B, and VEs and production capacities are similar to that of V-01 and NVSI-06-08 (i.e., 80% Vaccine A and 20% Vaccine B), respectively.

**Figure 1.**
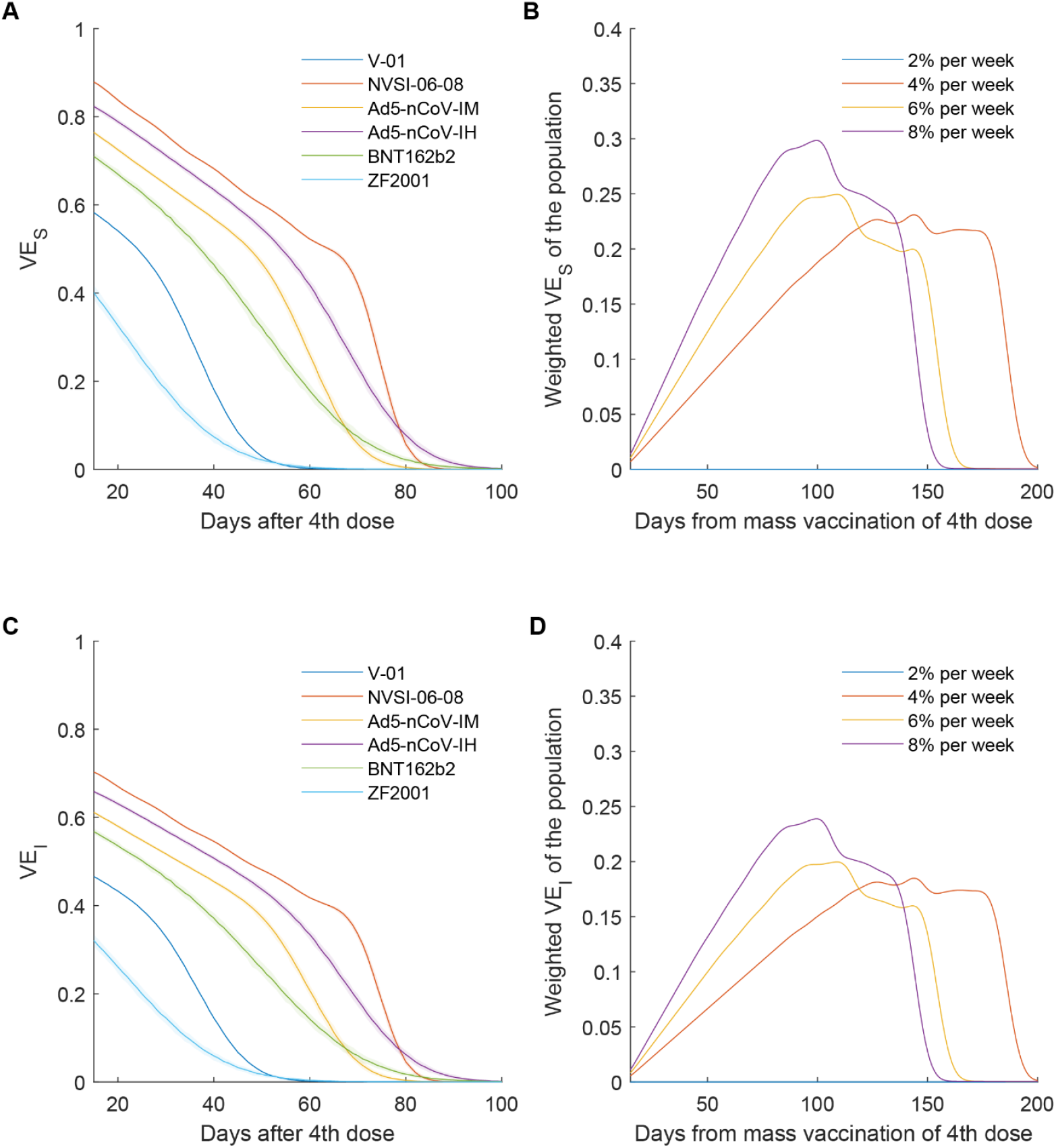
Vaccine effectiveness of heterologous boosting with the 4^th^ dose against Omicron BA.2 in reducing susceptibility (*VE*_*s*_) and infectiousness (*VE*_*I*_). (A-B) *VE*_*s*_ and *VE*_*I*_by days after the 4^th^ dose which is assumed to be equivalent to that conferred by a heterologous booster following 2 doses of inactivated virus vaccines (which was estimated from neutralising Ab titres 14-28 days after the 3^rd^ dose; **Table S1**). We assume that the Ab titres decay exponentially after the 3^rd^ and 4^th^ dose at the same rate (**Appendix**). (C-D) Population immunity against infection and infectiousness by days since the start of mass heterologous boosting at a maximum rate of vaccinating 2, 4, 6 and 8% of the population per week. Based on the national vaccination coverage data as of 28 November 2022, we assume the maximum vaccine uptake of the 3^rd^ and 4^th^ dose was 85%. We assume that 80% and 20% of vaccinees would be allocated with Vaccine A (with VEs similar to V-01) and Vaccine B (with VEs similar to NVSI-06-08), respectively. Assuming leaky vaccine action, we estimate population immunity as the product of vaccine uptake and VE. For example, if *VE*_*s*_ = 30% and vaccine uptake is 68%, then population immunity against infection is 21%.

### Effectiveness and availability of oral antivirals

Two oral antiviral medications, namely Molnupiravir and Nirmatrelvir/Ritonavir, have been used in Hong Kong since mid-March 2022 to treat COVID patients with a coverage of around 60% among eligible patients (i.e., patients aged 60 or above and high-risk patients under 60 years old). Given the current trajectory of COVID-19 antiviral availability and development in mainland China, we assume that Nirmatrelvir/Ritonavir would be used with 60% coverage when mainland China reopens, and that their effectiveness in reducing hospitalisations and deaths are similar to that observed in Hong Kong (**Table S3** and **Appendix**) ^11^.

### Effectiveness of PHSMs

Hong Kong has experienced six waves of community wide COVID transmission (with the fifth and sixth wave being Omicron) whereas Shanghai has had one driven by Omicron BA.2 that had led to 2 months of city-wide lockdown. We categorise PHSMs implemented during these waves into four levels and estimate their effectiveness from the associated changes in reproductive number (**Figure S1** and **Table S4**). In Hong Kong, PHSMs at Levels 1-3 reduced *R*_*t*_ by 15%, 47% and 55%, respectively. Level 4 PHSMs reduced *R*_*t*_ by 69% in Hong Kong and 72% in Shanghai.

We map mainland China’s combinations of PHSMs and their intensity to the abovementioned PHSM levels after the announcement of adjustment of COVID-19 response on 7 December 2022. We assume that the current PHSMs are as effective as Level 2, which would reduce *R*_*t*_ by 47%. A dynamic adjustment of PHSMs is anticipated during reopening, but we assume that PHSMs more stringent than

Level 4 would not be considered because it would lengthen the time needed for populations to attain their herd immunity thresholds.

### Modelling the spread and disease burden of Omicron

We assume that *R*_0_ of the SARS-CoV-2 variant that spread during reopening would be similar to that of the most recent prevailing strain worldwide, Omicron BA.5 (i.e., around 8.3) and 1,000 cases would be seeded into the reopened populations on the first ten days of reopening. With the vaccination coverage as of November 2022 as the starting point, we assess the respective and combined impact of vaccination, antivirals and PHSMs in five scenarios, and the effects of prioritising different age groups in four sub-scenarios **(Table 1)**. We adopt a previously used age-structured susceptible-exposed-infectious-removed (SEIR) meta-population model of SARS-CoV-2 transmission dynamics parameterized with local-specific age demographics and contact patterns ^12,13^. Population movements before, during and after *chunyun* among more than 300 prefecture-level cities are modelled based on previously constructed mobility networks adjusted with the relative changes in provincial transportation volumes between 2020 and 2021 ^14^.

**Table 1.** Scenarios of combining pharmaceutical and non-pharmaceutical interventions.

| Scenario | Combination of vaccinations, antivirals and PHSMs |
| --- | --- |
| **1** | • Status-quo with two-dose and three-dose vaccine uptake as of November 2022<br>• No 4<sup>th</sup>-dose vaccinations<br>• No antivirals<br>• No PHSMs |
| **2** | • Mass vaccination of 4<sup>th</sup> dose begins 30 days before the reopening and 4% of the population would receive the 4<sup>th</sup> dose per week. The maximum vaccine uptake of the 4<sup>th</sup> dose would reach 85% and 80% and 20% of individuals would receive Vaccine A and Vaccine B as the 4<sup>th</sup> dose.<br>• Antiviral coverage is 60%.<br>• No PHSMs |
| **3** | • Mass vaccination of 4<sup>th</sup> dose begins 30 days before the reopening and 4% of the population would receive the 4<sup>th</sup> dose per week. The maximum vaccine uptake of the 4<sup>th</sup> dose would reach 85% and 80% and 20% of individuals would receive Vaccine A and Vaccine B as the 4<sup>th</sup> dose.<br>• Antiviral coverage is 60%.<br>• PHSMs at Level 2 which reduce $R_t$ by 47% are implemented 14 days after the seeding of epidemics, and PHSMs at Level 2 are maintained between 15 and 74 days after the seeding of epidemics, and gradually relaxed between 75 and 104 days. |
| **4** | • Mass vaccination of 4<sup>th</sup> dose begins 30 days before the reopening and 6% of the population would receive the 4<sup>th</sup> dose per week. The maximum vaccine uptake of the 4<sup>th</sup> dose would reach 85% and 80% and 20% of individuals would receive Vaccine A and Vaccine B as the 4<sup>th</sup> dose.<br>• Antiviral coverage is 60%.<br>• PHSMs at Level 2 which reduce $R_t$ by 47% are implemented 14 days after the seeding of epidemics, and PHSMs at Level 2 are maintained between 15 and 74 days after the seeding of epidemics, and gradually relaxed between 75 and 104 days. |
| **5** | • Mass vaccination of 4<sup>th</sup> dose begins 30 days before the reopening and 6% of the population would receive the 4<sup>th</sup> dose per week. The maximum vaccine uptake of the 4<sup>th</sup> dose would reach 85%. Individuals would receive any of the five vaccines, including V-01, NVSI-06-08, Ad5-nCoV-IM, Ad5-nCoV-IH and BNT162b2, with equal probability as the 4<sup>th</sup> dose.<br>• Antiviral coverage is 60%.<br>• PHSMs at Level 2 which reduce $R_t$ by 47% are implemented 14 days after the seeding of epidemics, and PHSMs at Level 2 are maintained between 15 and 74 days after the seeding of epidemics, and gradually relaxed between 75 and 104 days. |

| Scenario | Vaccination coverage of the 4<sup>th</sup> dose and prioritisation of age groups in the vaccination programme under Scenario 4 |
| --- | --- |
| 4.1 | • The maximum vaccine uptake of the 4<sup>th</sup> dose would reach 85% in all age groups aged 3 or above.<br>• Roll-out of 4<sup>th</sup> dose vaccination follows the age-specific vaccine uptake of the 3<sup>rd</sup> dose as of November 2022. |
| 4.2 | • The maximum vaccine uptake of the 4<sup>th</sup> dose would reach 85% in all age groups aged 3 or above.<br>• Prioritise 4<sup>th</sup>-dose vaccination of older adults and seniors aged 60 or above. |
| 4.3 | • The maximum vaccine uptake of the 4<sup>th</sup> dose would reach 95% among 18-59 year olds and 85% in other age groups.<br>• Prioritise 4<sup>th</sup>-dose vaccination of adults aged 18 to 59 |
| 4.4 | • The maximum vaccine uptake of the 4<sup>th</sup> dose would reach 95% among 3-59 year olds and 85% in other age groups.<br>• Prioritise 4<sup>th</sup>-dose vaccination of adults aged 3 to 59 |

### Health system capacity

Among all Chinese cities, Hong Kong has one of the most well-resourced healthcare systems for managing the disease burden posed by COVID-19. As a best-case scenario, we use 100% and 200% of the maximum number of hospital beds designated for COVID-19 patients in Hong Kong in May 2022 (which correspond to its baseline/regular and surged capacity, respectively) as the benchmarks for health system capacity constraints across mainland China during reopening (**Table S3**). Assuming an average hospitalisation duration of 8 days ^15,16^, these constraints correspond to a daily incidence of 1.1 and 2.2 hospitalisations per 10,000 population, which also correspond to 21-42% of hospital beds in secondary/tertiary hospitals and 15-25% of all the existing hospital beds across all Chinese provinces (**Figure S2, Table S5** and **Table S6**).

### Reopening strategies

In view of the COVID-19 response adjustments announced on 11 November and 7 December, we assume that the full reopening would start in December 2022. Using simulations, we compare two reopening strategies:

**Strategy 1:** Simultaneously reopen all provinces on 1 December 2022 for simplicity of execution and to minimise the duration of the associated socio-economic disruption, including cities with major outbreaks recently (e.g., Beijing, Chongqing and Guangzhou)

**Strategy 2:** Start mass vaccination of the 3^rd^ dose homologous booster for the elderly and 4^th^ dose heterologous booster for other ages on 1 December 2022, and simultaneously reopen all provinces on 1 January 2023

Specifically, we simulate Strategies 1 and 2 in the following regions (**Figure S3**): (i) the greater Yangtze River Delta (YRD, including cities and counties in Shanghai, Jiangsu, Zhejiang, and Anhui) and greater Pearl River Delta (PRD, including cities and counties in Guangdong) as exemplars of Chinese megalopolises; (ii) Henan and Guangxi as exemplars of regions with a substantial proportion of rural populations.

## Results

### VE_s_ and population immunity conferred by 4^th^-dose heterologous boosting

We estimate that VE at 14 days after 4^th^-dose heterologous boosting in reducing Omicron susceptibility (*VE*_*s*_), infectiousness (*VE*_*I*_), hospitalisations (*VE*_*H*_) and deaths (*VE*_*D*_) are: (i) 58%, 47%, 96% and 98% for Vaccine A; and (ii) 88%, 70%, 96% and 98% for Vaccine B, respectively (**Table 2**). However, *VE*_*s*_ and *VE*_*I*_wane quickly such that both vaccines would have minimal effect in reducing Omicron transmission 60-90 days after 4^th^-dose boosting (**Figure 1**). The population immunity against Omicron transmission during reopening is sensitive to the start time of 4^th^-dose boosting and the associated rollout rate (with respect to the time of reopening). Assuming that 4^th^-dose uptake would reach 85% at a weekly rate of 8%, 6% and 4% of the population, population immunity against infection and infectiousness would reach 20-30% and 15-25% within 70, 90, and 120 days, respectively. Slower rollout of the 4^th^ dose would fail to generate significant population immunity against Omicron transmission (**Figure 1**). Immunity from 4^th^-dose boosting would be sustained at peak levels for about 60 days and then drop substantially afterwards. Taken together, if population immunity is one of the prerequisites for safer reopening, 4^th^-dose heterologous boosting should start and finish within 1-2 months before the commencement of reopening by vaccinating 4-8% of the population per week.

**Table 2.**
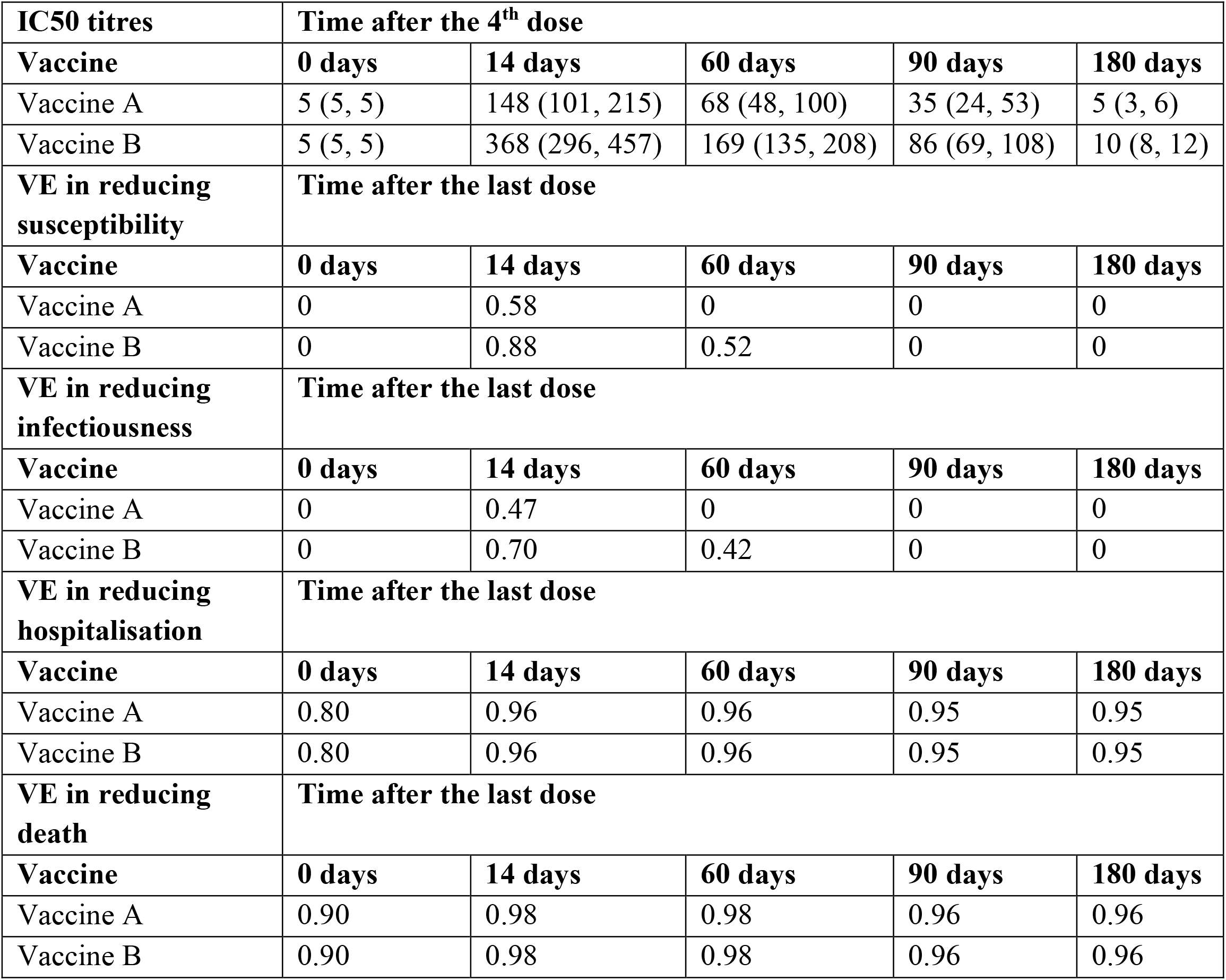
Assumption about neutralising antibody titres (IC50) and vaccine effectiveness (VE) against Omicron BA.2 after the fourth dose in mainland China (with BBIBP-CorV as the first three doses)

### Impact of combining vaccination, antiviral treatment and PHSMs

We first consider status quo without 4^th^-dose vaccination, antivirals and PHSMs (**Scenario 1** in **Table 1**) and epidemics are simultaneously seeded in all provinces. In this case, reopening at the status quo would result in a cumulative mortality burden of 684 per million (**Figure 2**).

**Figure 2.**
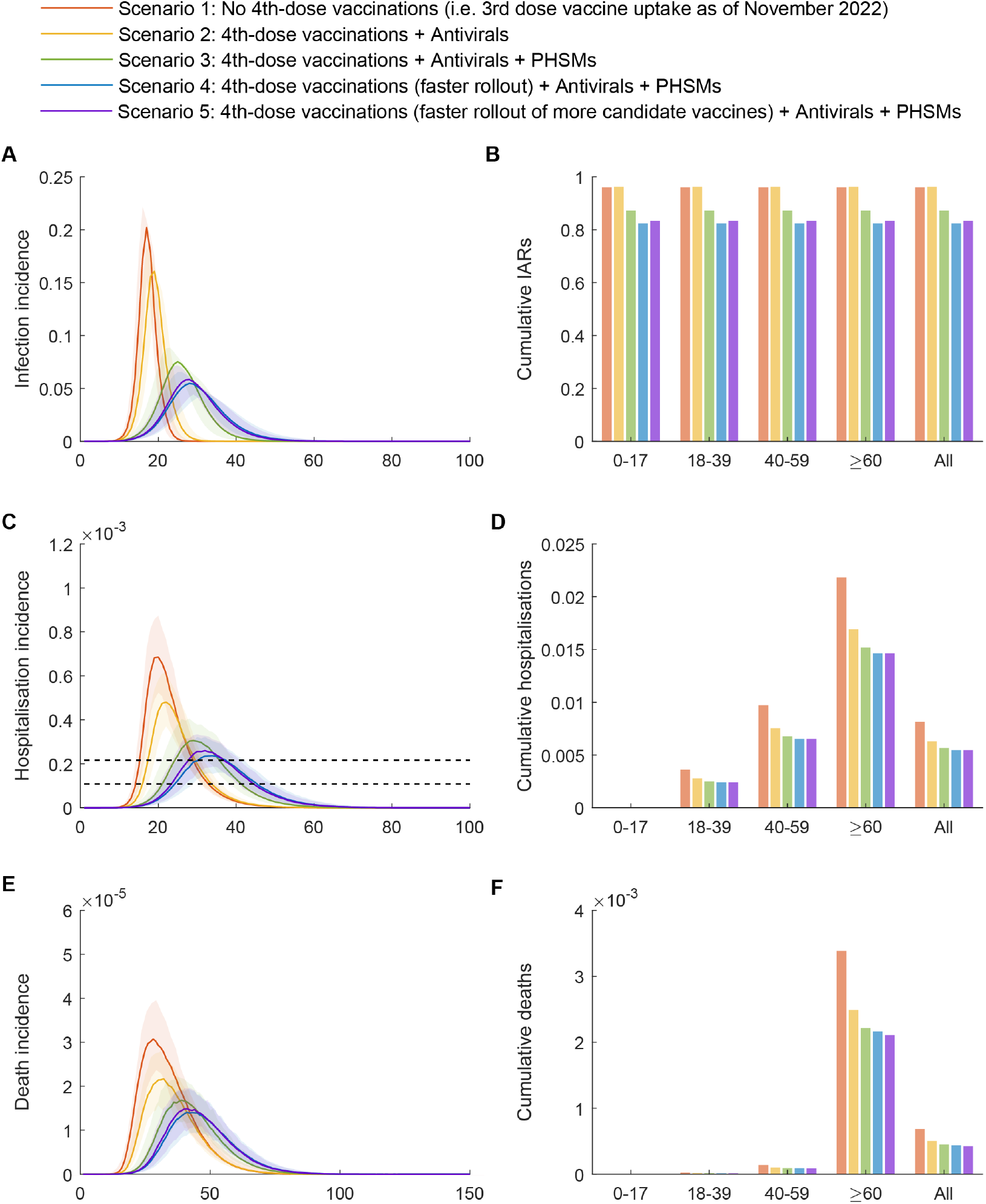
The impacts of 4^th^ dose heterologous boosting, antiviral treatments and PHSMs if they are combined during reopening. Please see Table 1 for descriptions of the five scenarios. **(A) Infection incidence as proportion of the population. (B) Cumulative infection attack rates by age. (C-D) Daily and cumulative number of cases requiring who require hospitalisation. (E-F) Daily and cumulative incidence of death**. Solid lines indicate the 50th percentile of the estimates and shades indicate the ranges between 2.5th and 97.5th percentiles. The two horizontal dashed lines indicate the two health system capacity constraints based on the experience of Hong Kong during its Omicron wave (see Methods).

We next consider vaccinations, antiviral treatments and PHSMs individually (**Figure S4**). Assuming that the basic reproductive number *R*_0_ of Omicron BA.5 is 8.3 (7.8-8.9) and a fast rollout of the 4th dose would provide 25% and 20% population immunity against infection and infectiousness (**Figure 1**), *R*_*t*_ of Omicron BA.5 under Leve1 1-4 PHSMs would be around 4.2, 2.6, 2.2 and 1.5, respectively. If vaccination, antiviral treatments and PHSMs are only implemented in isolation, the disease burden posed by reopening would substantially exceed the health system capacity in all provinces.

Therefore, we consider vaccination, antiviral treatments and PHSMs in different combinations **(Table 1). Scenario 2** shows that with vaccination and antivirals but not PHSMs, hospitalisations and deaths would again exceed by a large margin the capacity of local health systems for 2-3 weeks in all provinces (**Figure 2**). Specifically, the daily number of hospitalisations would peak at 4.8 (3.8-6.2) per 10,000 population which is more than twice the surge capacity constraint (i.e., 2.2 per 10,000), although the cumulative number of deaths would be reduced by 26% to 503 per million compared with Scenario 1.

Similarly, **Scenario 3** shows that, although adding PHSMs could further reduce the spread and overall mortality (to 448 per million), peak hospitalisation burden would still overload the health systems for weeks albeit with smaller excess margins. **Scenarios 4 and 5** show that in addition to PHSMs, faster rollout of 4^th^-dose boosting would slightly reduce the overall mortality to 426 and 416 per million, but they would be unable to completely avert overloading of the local health systems beyond the surge capacity constraint.

Increasing booster uptake to 95% among 18-59 and 3-59 years old would further reduce the overall mortality to 305 and 249 per million, respectively (**Figure 3**). However, prioritising boosting among younger age groups that contribute more to transmission would only modestly reduce the peak incidence of infection and hospitalisation with minimal additional indirect protection of the elderly population. Peak hospitalisation incidence would still exceed the baseline capacity constraint (i.e., 1.1 per 10,000) even when 4^th^-dose uptake reaches 95% among 3-59 years old and >85% in other age groups.

**Figure 3.**
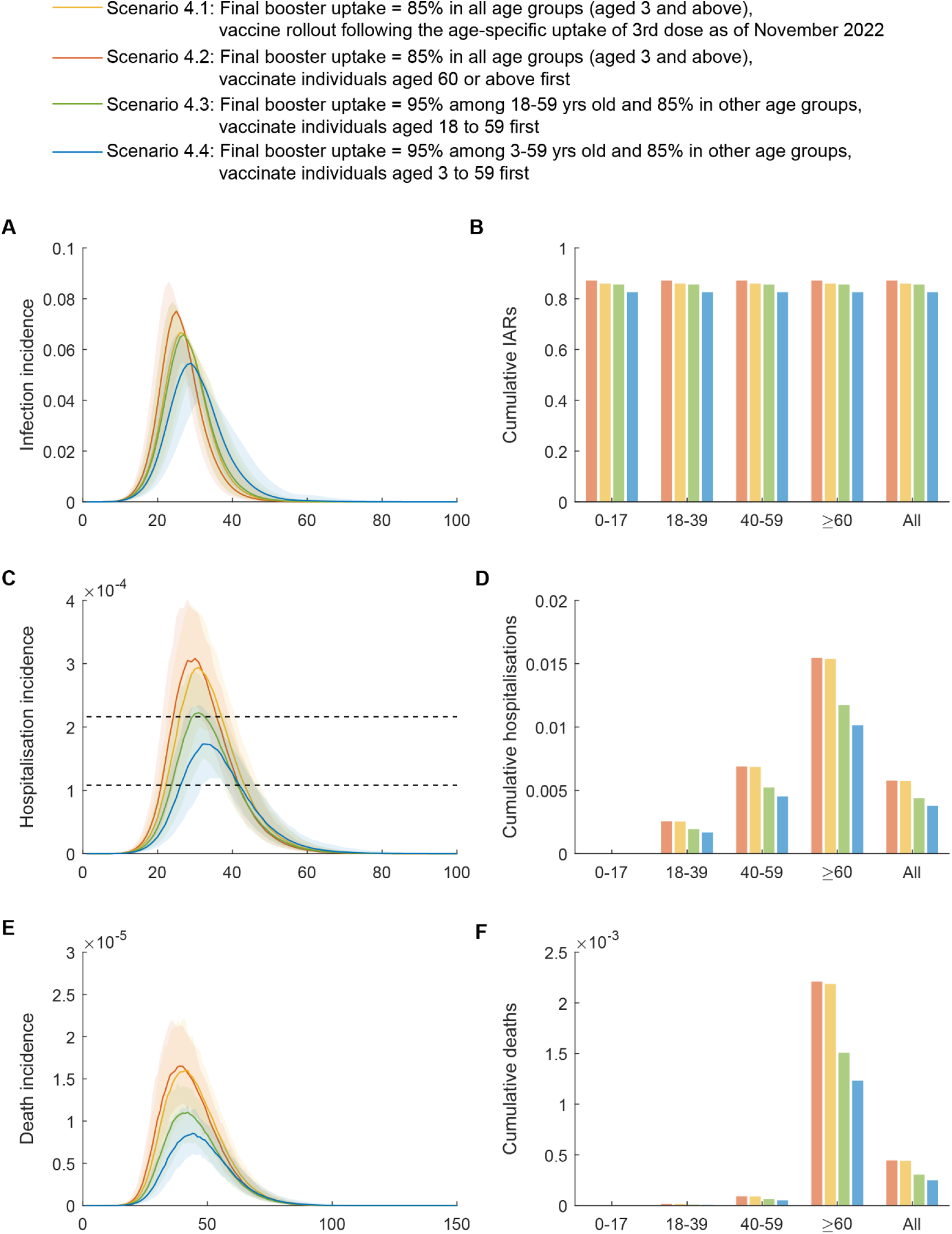
The impact of prioritising different age groups for 4^th^ dose heterologous boosting. We consider four prioritisation schemes for boosting (Table 1). Other parameters are the same as Scenario 4 in **Figure 2**.

Populations with 4^th^-dose uptake below 85% would require more stringent PHSMs to reduce *R*_*t*_ by >65% (entailing closure of most indoor premises and different sectors of business, amongst others) in order to prevent peak daily incidence of hospitalisation from exceeding 2.2 per 10,000 (**Table S5**). Populations with older age structures also require more stringent or prolonged PHSMs for safe reopening, because those aged 60 or above have much higher risks of hospitalisation and death if infected. In what follows, we only simulate **Scenario 4** unless specified otherwise.

### Simultaneous reopening of all provinces without 4^th^ dose heterologous booster (Strategy 1)

Under **Strategy 1**, simultaneous reopening of all provinces without 4^th^ dose vaccination would result in almost synchronised epidemics in both YRD and PRD, especially under their higher connectivity with all provinces, cities and counties during the *chunyun* period (**Figure 4**).

**Figure 4.**
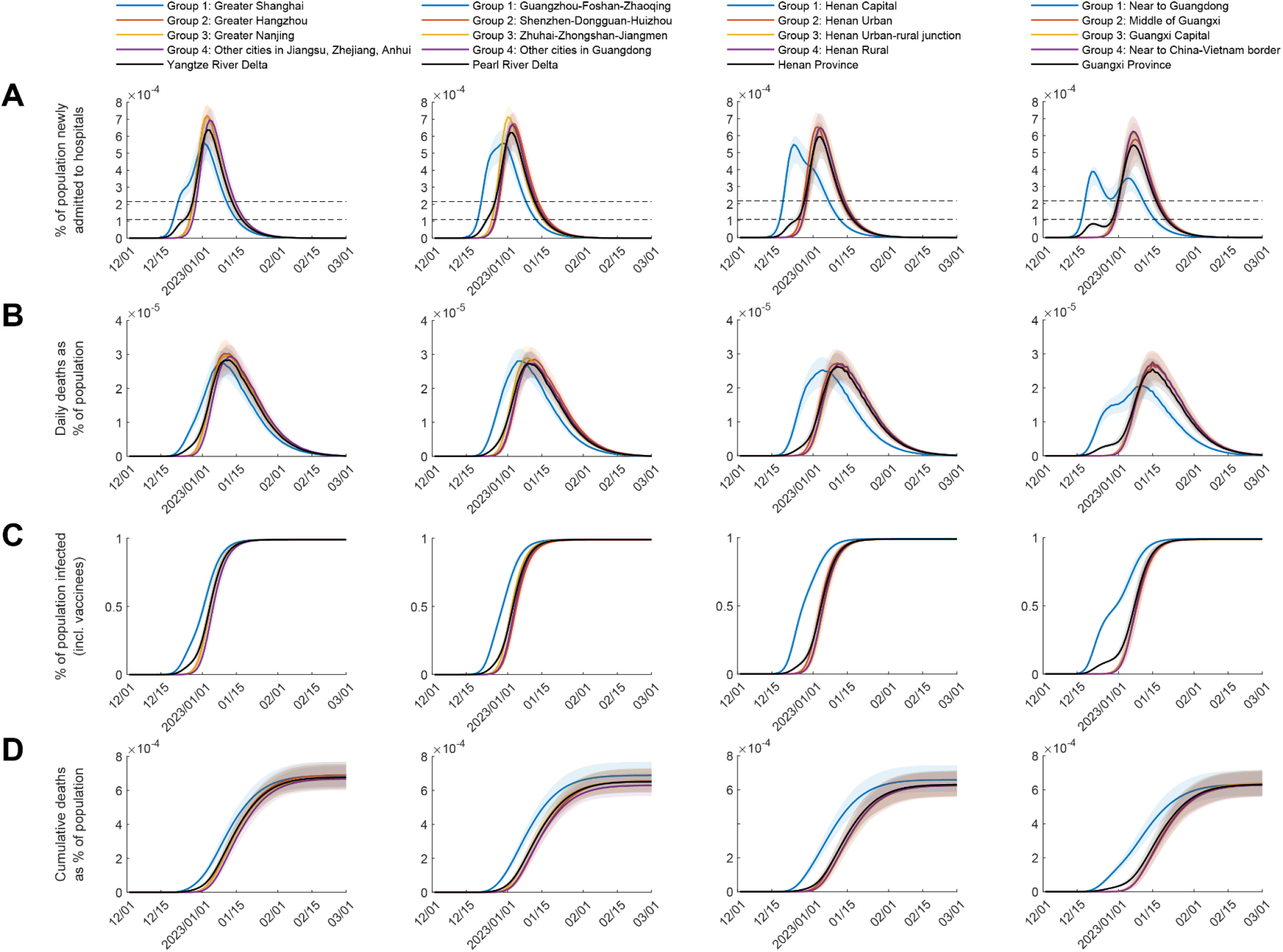
Simultaneous reopening of Yangtze River Delta Region, Pearl River Delta Region, Henan, and Guangxi without 4^th^ dose vaccination. We assume that mass vaccination of the 4^th^ dose would not be implemented. Antiviral coverage is 60%. PHSMs at Level 2 are implemented 14 days after the seeding of epidemics, and PHSMs at Level 2 are maintained between 15 and 74 days after the seeding of epidemics, and gradually relaxed between 75 and 104 days. In four regions, we assume COVID-19 outbreaks are seeded by importations in Group 1 (Shanghai, Guangzhou, Zhengzhou, and Guilin) on 1 December 2022. Solid lines show the 50^th^ percentile of the estimations and shades show the ranges between 10^th^ and 90^th^ percentiles. Other parameters are the same as Scenario 4 in **Figure 2**.

The epidemic lasts longer and peaks slightly earlier in the cities where epidemics are first seeded. In other cities within YRD and PRD, daily incidence of hospitalisation peaks at nearly the same time, and the surge in hospital admissions would exceed the capacity of health systems in all cities. At peak, the daily number of hospitalisations could reach 6-7 per 10,000 population which is about 3-4 times the surge capacity threshold of health system capacity (2.2 hospitalisations per 10,000). The mean daily incidence of deaths is 2.3-3.5 per 100,000 at peak, and the mean cumulative incidence of deaths is 568-770 per million in all populations. For example, the peak daily number of deaths is 573-872 and the cumulative number of deaths is 14,138-19,166 in Shanghai (24.89 million population).

The epidemics in Henan and Guangxi would be similar to that in YRD and PRD except for a longer time lag between the first epidemic wave in cities where epidemics are seeded and the subsequent epidemics in other cities (which are synchronised) due to relatively weaker population mobility between them (**Figure 4**).

### Simultaneous reopening of all provinces with 4^th^ dose heterologous booster (Strategy 2)

Under **Strategy 2**, simultaneous reopening of all provinces with 4^th^ dose vaccination would similarly result in almost synchronised epidemics in both YRD and PRD, but mass vaccination with 4^th^ dose heterologous boosting would reduce the peak incidence of hospitalisations and deaths (**Figure 5**). At peak, the daily number of hospitalisations is reduced to 4-6 per 10,000 population which is about 1.5-2.5 times the surge capacity threshold of health system capacity (2.2 hospitalisations per 10,000). The mean daily incidence of deaths is also reduced to 1.7-2.4 per 100,000 at peak, and the mean cumulative incidence of deaths is 434-615 per million in all populations. For example, the peak daily number of deaths is 424-598 and the cumulative number of deaths is 10,803-14,885 in Shanghai.

**Figure 5.**
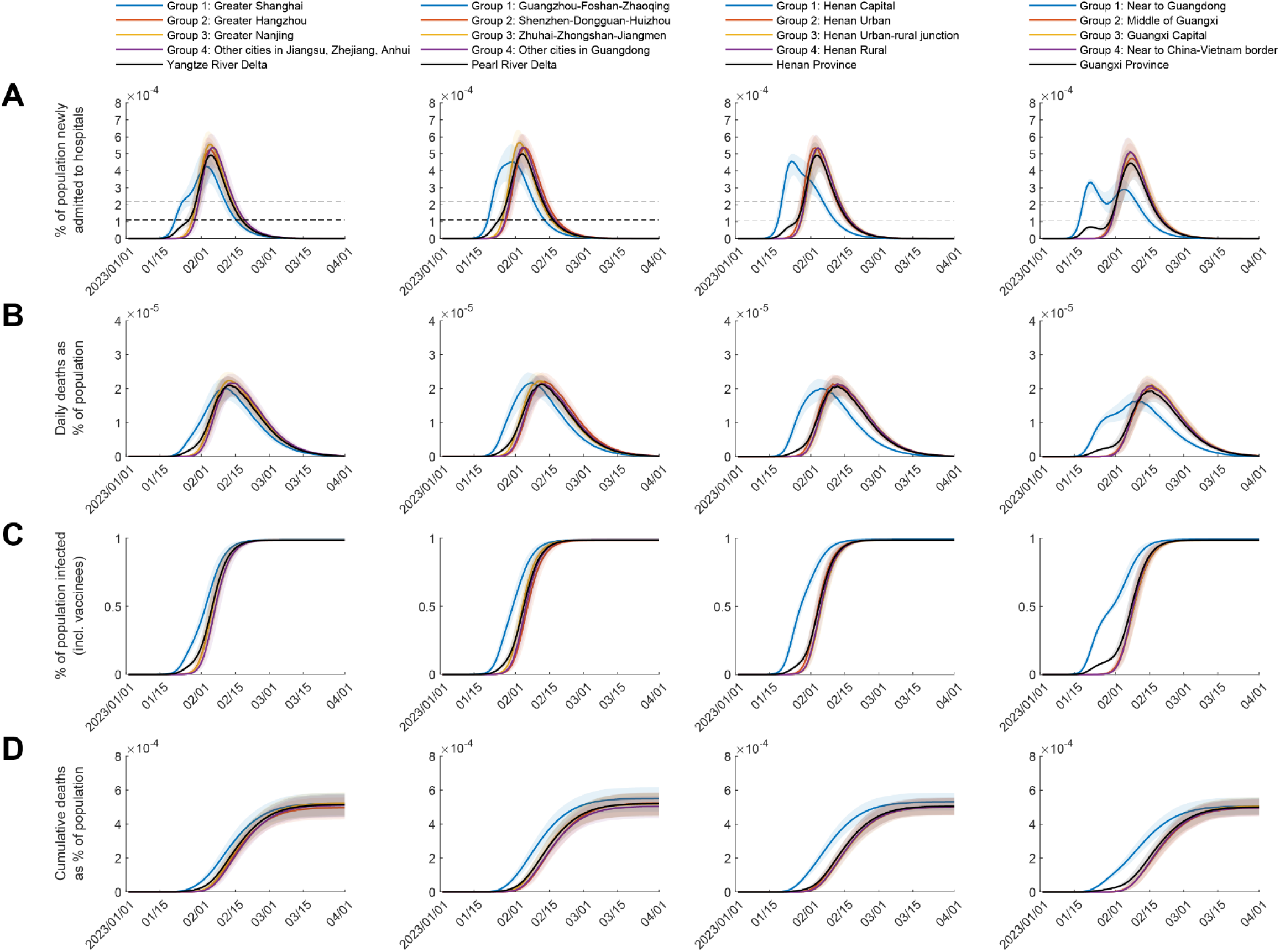
Simultaneous reopening of Yangtze River Delta Region, Pearl River Delta Region, Henan, and Guangxi with 4^th^ dose vaccination. We assume that mass vaccination of the 4^th^ dose starts 30 days before the reopening to vaccinate 6% of the population per week, and uptake of the 4^th^ dose would reach 85%. Antiviral coverage is 60%. PHSMs at Level 2 are implemented 14 days after the seeding of epidemics, and PHSMs at Level 2 are maintained between 15 and 74 days after the seeding of epidemics, and gradually relaxed between 75 and 104 days. In four regions, we assume COVID-19 outbreaks are seeded by importations in Group 1 (Shanghai, Guangzhou, Zhengzhou, and Guilin) on 1 January 2022. Solid lines show the 50^th^ percentile of the estimations and shades show the ranges between 10^th^ and 90^th^ percentiles. Other parameters are the same as Scenario 4 in **Figure 2**.

### More stringent PHSMs are required during reopening

Given that the surge of COVID-19 hospitalisations would overload health systems of all provinces in Strategies 1-2, we estimate that more stringent PHSMs are required during reopening (**Figure 6**). Without 4^th^ dose heterologous boosting, implementing PHSMs at Level 4 could only reduce the peak hospitalisation to 4.3 per 100,000. Therefore, more stringent PHSMs to reduce *R*_*t*_ by 80-88% (similar to lockdown) are required to keep peak hospitalisation below the surge capacity threshold of health system. There is a trade-off between higher vaccination coverage and less stringent PHSMs: if 4^th^ dose heterologous vaccination coverage could reach >75% or above nationally, PHSMs at Level 3 would be able to keep peak hospitalisation below the surge capacity threshold of health system.

**Figure 6.**
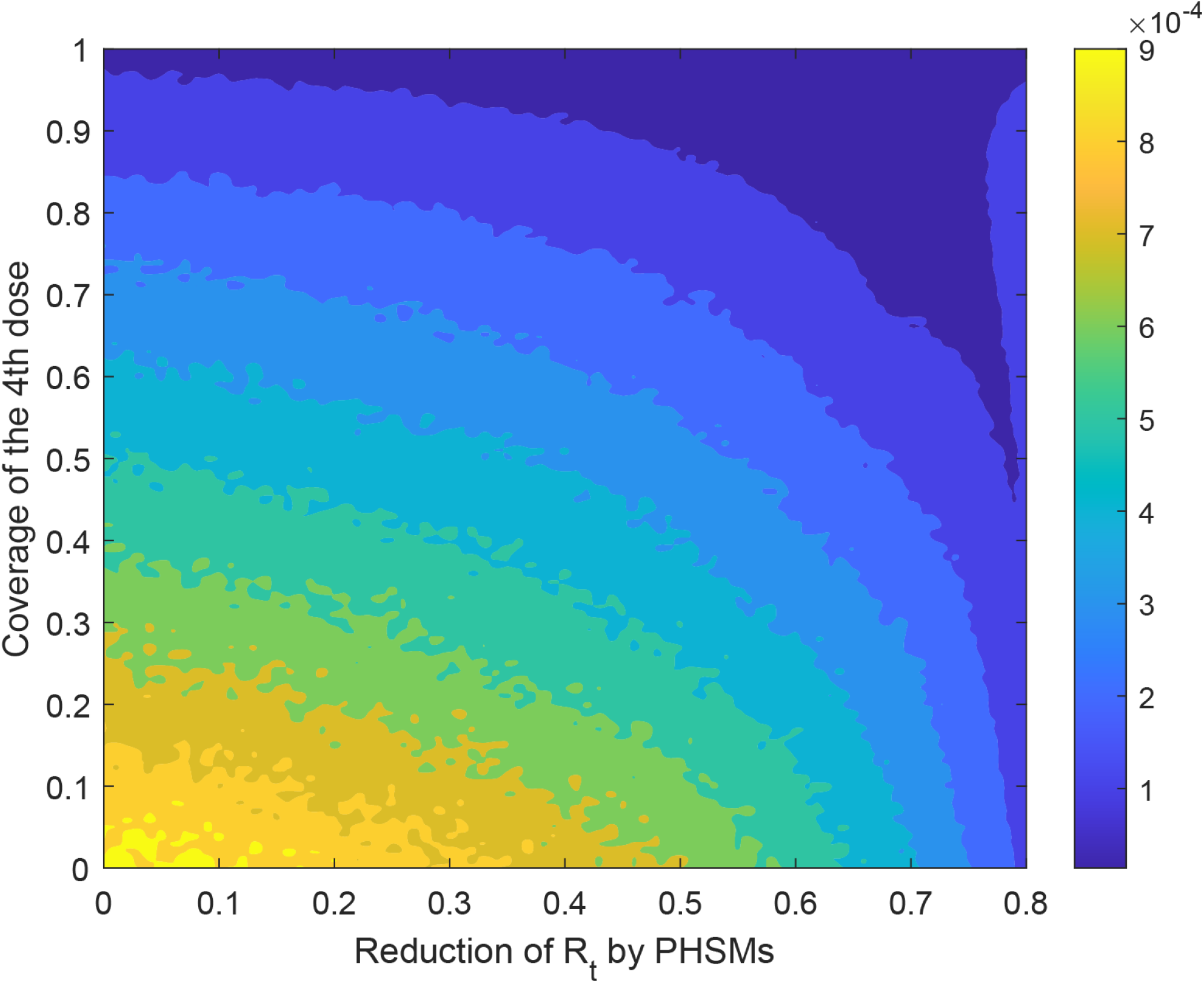
Peak daily incidence of hospitalisation as a function of PHSM intensity and 4^th^ dose uptake. We assume that reopening would begin 30 days after the start of 4^th^-dose boosting and 6% of the population would receive the 4^th^ dose per week. PHSMs would be implemented 14 days after the seeding of epidemics and maintained for 60 days (i.e., until day 74 after reopening has begun). PHSMs would then be gradually over the next 30 days. Other parameters are the same as Scenario 4 in **Figure 2**.

## Discussion

Our results suggest that local health systems across all provinces would be unable to cope with the surge of COVID-19 cases posed by reopening in December 2022 – January 2023 in the context of the adjusted COVID-19 responses announced on 7 December ^4^. However, safer exit from dynamic zero-COVID could be achieved by adopting a multi-pronged approach comprising vaccination, antiviral treatment, PHSMs and sequential reopening. As of 28 November 2022, the nationwide three-dose vaccine uptake was 57% and 69% among population aged <60 and ≥60. As such, it is crucial to substantially boost population immunity to minimise morbidity and mortality during the reopening.

A high uptake of heterologous boosting with an efficacious fourth dose is the cornerstone for reopening more safely. As of 7 December 2022, among all domestic vaccines under emergency use or development, recombinant protein subunit vaccines (e.g., V-01, SCB-2019 and SCTV-01C) and inhaled adenovirus-vectored vaccines (e.g., Ad5-nCoV-IH) have shown good potential for inducing immunity against Omicron infections (**Table S1**). Boosting population immunity (especially among the elderly population and high-risk groups) are current top priorities for setting the stage for safe reopening. Increasing vaccine uptake in all age groups, including children aged under 3, should be also considered. Given the fast waning of VEs, mass vaccination programme of the fourth dose should start within 30-60 days before reopening and complete within 60 days (**Figure 1**).

China is a highly heterogenous country with megalopolises and developed urban areas in the East and Southeast, but the majority of rural areas are in the North, Northwest, and Southwest. Economically prosperous regions have relatively more abundant healthcare resources and are therefore more resilient in coping with the surge of cases and hospitalisations during reopening (**Table S5**). When these regions reopen, PHSMs should ideally be kept at Level 2-3 to minimise the associated socio-economic disruption whenever possible, but PHSMs at Level 4 should be considered when surge of hospitalisations is expected to overload the health system. Antivirals should be prescribed in a timely manner in symptomatic cases at high coverage, and treatment and management of COVID-19 patients should be risk stratified. For example, a primary-care supported home recovery programme could be introduced for low-risk patients (i.e., those who are asymptomatic or mildly symptomatic), whereas high-risk patients should be provided with necessary medical care (especially with timely antivirals) and closely monitored with the help of telemedicine. In this regard, antivirals should be made more widely accessible at a much lower cost.

In contrast, a large proportion of China’s elderly population live in rural areas where treatment and care are most needed during the reopening transition. Therefore, ramping up temporal hospital capacity in rural areas should be emphasised in the interim. Ideally, vaccination coverage of the 3^rd^ and 4^th^ dose should exceed 90%, and logistics infrastructure should be set up for efficient and timely on-demand antiviral distribution at the household level. When these rural areas reopen, more stringent PHSMs of Level 4 could be considered near the epidemic peaks to further minimise the surge demand of hospital care. However, in the context of reopening, the functional aim of PHSMs is to strike an optimal balance between: (i) keeping the peak hospital load just below the surge capacity constraint **(Figure 6)**; and (ii) allowing epidemics to infect around 1-1/*R*_0_ (the herd immunity threshold) proportion of the population within 6 months in order for natural infections to generate enough long-lasting hybrid immunity against substantial resurgence afterwards. As such, PHSMs should not be overly stringent and city-wide lockdown (i.e., PHSMs above Level 4) should be avoided, because cases will resurge as soon as those PHSMs are lifted otherwise.

Although safe reopening might reduce the surge of COVID-19 severe cases to more manageable levels, reopening mainland China at an initial *R*_*t*_ close to 3 would still result in a large number of infections that could potentially accelerate mutation, selection and evolution of SARS-CoV-2 viruses ^17,18^. Genomic surveillance for SARS-CoV-2 variants must be strengthened, particularly in megalopolises that are highly connected domestically and internationally. Multidisciplinary studies of the transmissibility, severity, immune/vaccine escape of variants and antiviral resistance should be coordinated nationwide to track the evolution of the pandemic ^19^.

In conclusion, although the surge of disease burden posed by reopening in December 2022 – January 2023 would likely overload most local health systems nationwide, a reopening strategy that combines vaccination, antiviral treatment and PHSMs could allow China to exit zero-COVID more safely. This would require a nationally coordinated effort for reopening, including planning, execution, and surveillance.

## Data Availability

We collated all data from publicly available data sources. All data included in the analyses are available in the main text or the supplementary materials.

## Contributors

All authors designed the study, developed the model, analysed data, interpreted the results, and wrote the manuscript.

## Declaration of interests

The authors declare no competing interests.

## Funding

This research was supported by the COVID-19 Vaccines Evaluation Program (COVEP) of The Chinese Center for Disease Control and Prevention (grant no.: 2202-HK-25), Health and Medical Research Fund (grant no.: 21200122, CID-HKU2 and COVID19F05), Health and Medical Research Fund Research Fellowship Scheme (grant no.: 06200097), General Research Fund (grant no.: 17110020), and the AIR@InnoHK administered by Innovation and Technology Commission of The Government of the Hong Kong Special Administrative Region. KL was supported by the Enhanced New Staff Start-up Research Grant from LKS Faculty of Medicine, The University of Hong Kong. The funders of the study had no role in study design, data collection, data analysis, data interpretation, or writing of the report. The corresponding author had full access to all the data in the study and had final responsibility for the decision to submit for publication.

## Appendix

### Estimating vaccine effectiveness of heterologous boosting in reducing Omicron susceptibility and infectiousness

As of 28 November 2022, the two- and three-dose vaccine uptake in mainland China was 91% and 57%, respectively. The corresponding uptake among those aged 60 or above was 86% and 69%, respectively (http://www.nhc.gov.cn/).

Since more than 95% of vaccines administered in mainland China currently are inactivated virus vaccines, we assume that all vaccinees would receive inactivated virus vaccines (namely, BBIBP-CorV or CoronaVac) for the first three doses. The IC50 neutralising antibody (Ab) titres against Omicron after the first three doses were obtained from the Brazilian study RHH-001 ^6^ and the Phase-3 trial of V-01 vaccine ^20^.

We assume that 4^th^-dose heterologous boosting would be delivered via two hypothetical vaccines A and B whose VEs and production capacities are similar to that of V-01 and NVSI-06-08 (**Table 2, Table S1 and Table S2**):

1. V-01 is a recombinant SARS-CoV-2 fusion protein vaccine (i.e. RBD dimer-IFN-Pan Fc fusion protein) developed by Livzon. In the Phase-3 trial of 10,218 participants in Pakistan and Malaysia with a follow-up period of 60-90 days, the vaccine efficacy of V-01 as the third dose was 64% (23-83) and 39% (3-62) against Omicron infection among 4,935 participants who had received BBIBP-CorV or CoronaVac vaccines in their primary course ^20^.
2. NVSI-06-08 is a recombinant COVID-19 vaccine based on the antigen of a mutation-integrated trimetric RBD developed by Sinopharm. In the Phase-2 trial, the neutralising Ab titres against Omicron was increased to 368 (95% CI: 296-457) among participants who had received BBIBP-CorV as their primary course and NVSI-06-08 as the third dose ^21^.

We apply the Khoury method to predict the vaccine effectiveness (VE) in reducing Omicron susceptibility and infectiousness as described in Leung et al ^9^. The prediction is based on the neutralising Ab data from Brazil and Hong Kong ^6,22,23^ and the VE data from Phase-3 trial of V-01, RHH-01 study from Brazil, and UKHSA reports from the UK ^6,20,24^. Using the same method, we model the waning of VE based on the following assumptions regarding the waning of Ab titres: (i) The waning rate of Ab titres after the third dose is the same for inactivated virus vaccines and BNT162b2; (ii) the waning rate after the third and fourth dose are the same. We estimate the waning rate of Ab titres for BNT162b2 by mapping the neutralising titres after three doses of BNT162b2 vaccines from Cheng et al ^22^ to the 3-dose VEs estimates from UKHSA by Andrews et al ^24^.

It is believed that the immune response after receiving the third or fourth dose of vaccine, especially cellular immunity (e.g., via T cells), may provide greater and more long-lasting protection against severe disease than mild or asymptomatic infection ^25-29^. As such, we assume that the VE against severe disease, hospitalisations and death after 4^th^-dose heterologous boosting would be similar to the corresponding VEs among recipients of three doses of CoronaVac vaccines in Hong Kong ^10^, regardless of VE waning over time and the emergence of VOCs (**Table 2**).

#### Estimating the vaccine-induced population immunity

We model the rollout of vaccination programme under the following assumptions:

1. The target vaccine uptake of the 2^nd^, 3^rd^ and 4^th^ dose is 90%, 85% and 85% for all age groups, respectively, based on the age-specific 2-dose and 3-dose vaccine uptake in mainland China as of 28 November 2022.
2. For the 4^th^ dose, we assume that 80% of vaccinees would be allocated with Vaccine A, and the remaining 20% of vaccinees would be allocated with Vaccine B. According to the press release of V-01 vaccine manufacturer Livzon on 12 April 2022, their Guangdong branches have been approved to produce the V-01 vaccine. The annual production capacity could reach 1.5 billion doses of packaged V-01 vaccines (https://www.livzon.com.cn/ and http://www.news.cn/english/2021-08/30/c_1310155999.htm). According to the press release of NVSI-06-08 vaccine manufacturer Sinopharm on 3 April 2022, their factories in Beijing and Gansu have been producing NVSI-06-08 vaccines. Between October and December 2021, Sinopharm had produced 80 million doses of NVSI-06-08 vaccines and delivered 20 million doses outside mainland China (http://www.sinopharm.com/s/1223-4131-40132.html). Currently, the annual production capacities of the two vaccines are 1.5 billion and 0.32 billion doses and we assume a similar ratio in the vaccine allocation of the 4^th^ dose.
3. The roll-out of the 4^th^ dose will be considered after the 3^rd^-dose uptake has reached 80%.
4. The time interval between the 1^st^ and 2^nd^ dose is 28 days.
5. The time interval between the 2^nd^ and 3^rd^ dose is at least 90 days.
6. The time interval between the 3^rd^ and 4^th^ dose is at least 180 days.
7. The maximum weekly vaccination rate is 8% of the population. During the rapid rollout of mass vaccination in 2021, the maximum number of vaccine doses given to the Chinese population reached 24 million per day in May 2021, which was equivalent to a weekly rate of 12% of the population (http://www.nhc.gov.cn/).

#### Estimating the effects of PHSMs from the past waves of COVID-19 in Hong Kong and Shanghai

Since the emergence of the COVID-19 pandemic, various PHSMs have been used to suppress and mitigate the spread of SARS-CoV-2 in Hong Kong. We analyse data on locally laboratory-confirmed cases of the first four waves of COVID-19 outbreaks and estimated the daily effective reproductive number (*R*_*t*_) to estimate the changes in transmissibility over time (**Figure S1**). During each wave, PHSMs were progressively tightened with the increase of reported cases. Without loss of generality, we group the PHSMs into four levels in each wave using the time when civil servants were required to work from home (WFH) as a cut-off:

1. **Level 1:** Voluntarily universal face masking and improved hand hygiene
2. **Level 2: Level 1** PHSMs **+** PHSMs announced or implemented before civil servants WFH, which usually included tightened social distancing measures in restaurants and indoor leisure facilities, and closure of kindergartens and primary schools of Grade 1-3 or Grade 1-4.
3. **Level 3: Level 2** PHSMs **+** PHSMs announced or implemented together with civil servants WFH, which often included closure of most indoor leisure facilities, closure of all schools, no dine-in in restaurants after 9 pm.
4. **Level 4: Level 3** PHSMs **+** PHSMs announced or implemented after civil servants WFH, which included more stringent control measures of restaurants, such as no dine-in after 6 pm or all day.

We assume that the basic reproductive number was 2.0 for the first wave (the original virus strain), 2.2 for the second wave (70% of original virus and 30% of D614G mutant virus which was estimated to be 30% more transmissible than the original virus), 2.6 for third and fourth wave (D614G mutant virus), in the absence of any PHSMs and COVID-19 vaccination. Then we estimate the effectiveness of PHSMs at Level 1-4 by overlaying these PHSMs with *R*_*t*_ in the first four waves (**Figure S1 and Table S4**).

In the fifth wave of Omicron BA.2 in Hong Kong, we estimate the effectiveness of Level 4 PHSMs in parameter inference in the epidemic model ^16^. Similarly, we estimate the effectiveness of PHSMs in Shanghai before the lockdowns of Pudong (the east of Huangpu River) on 27 March 2022 and Puxi (the west of Huangpu River) on 1 April 2022.

#### Reopening in regions with substantial proportion of rural populations

We define urban, urban-rural junction and rural area as administrative regions with <64%, 64-71% and >71% of sub-administrative regions coded as “rural”, based on the coding of National Bureau of Statistics (http://www.stats.gov.cn/tjsj/tjbz/tjyqhdmhcxhfdm/2020/index.html). The urbanisation rate of China is 60% in 2019, and at the national level, the 20^th^, 40^th^, 60^th^ and 80^th^ percentile of the proportion of sub-administrative regions coded as “rural” are 55%, 64%, 71% and 77%, respectively.

In Henan, we assume that the reopening starts simultaneously with the epidemics seeded in the provincial capital region (Zhengzhou and Xuchang) and spread to urban area, urban-rural junction, and rural area, respectively (**Table S7**). In Guangxi, we assume that the reopening starts with the epidemics seeded in the area adjacent to PRD/Guangdong and spread geographically in a diffusion manner to the middle of Guangxi, provincial capital, and area next to China-Vietnam border (**Figure S3**).

#### Impact of vaccination, antiviral treatment and PHSMs if they are singly implemented during reopening

We assess the impact of vaccination, antiviral treatments and PHSMs if they are singly implemented during reopening as follows: (i) 4^th^-dose boosting would be rolled out at a weekly rate of 6% of population; (ii) antiviral treatments would be deployed at 60% coverage; and (iii) Level 4 PHSMs (which reduce *R*_*t*_ by 69-72%; see **Table S4**) are implemented between day 15 and 74 of reopening and then gradually relaxed to normalcy by Day 104. The final cumulative infection attack rate would be >95%, >95% and 79% in the three scenarios, respectively. The number of cases who require hospitalisation would far exceed the local health system capacity for weeks to months with cumulative incidence of 91, 266, 323 per 10,000 and cumulative deaths of 8.1, 23.5 and 33.9 per 10,000, respectively (**Figure S4**). Among those aged 60 or above, the cumulative number of hospitalisations is 242, 714 and 868 per 10,000 and the cumulative number of deaths is 40, 116 and 168 per 10,000. That is, this age group accounts for more than 50% of hospitalisations and 92% of deaths associated with COVID-19.

**Table S1.** Neutralising antibody titres (IC50 or PRNT50 or PNAb50) of COVID-19 vaccines against Omicron by time and dose.

| Vaccine (combination) | Time since the last dose |  |  | Source |
| --- | --- | --- | --- | --- |
|  | 0 days | 14/28 days | 180 days |  |
| CoronaVac × 2 | -- | 11 (9, 15) | 11 (9, 15) | Costa Clemens et al <sup>6</sup> |
| CoronaVac × 2 + BNT162b2 | 10 (10, 10) | 223 (108, 458) | -- |  |
| CoronaVac × 2 + Ad26.COV2-S | 10 (10, 10) | 138 (72, 264) | -- |  |
| CoronaVac × 2 + ChAdOx1 | 10 (10, 10) | 102 (57, 182) | -- |  |
| CoronaVac × 3 | 11 (9, 15) | 17 (11, 26) | -- |  |
| BBIBP-CorV × 2 | -- | 6 (5, 8) | -- | Ai et al <sup>30</sup> |
| BBIBP-CorV × 2 + ZF2001* | 19 (11, 32) | 109 (54, 221) | -- |  |
| BBIBP-CorV × 3 | 5 (5, 5) | 48 (26, 84) | -- |  |
| CoronaVac × 2 + Ad5-nCoV-IM | -- | 261 (178, 382) | -- | Li et al <sup>31</sup><br>Zhang et al <sup>32</sup> |
| CoronaVac × 2 + Ad5-nCoV-IH | -- | 320 (191, 538) | -- |  |
| CoronaVac × 2 + ZF2001* | -- | 86 (59, 127) | -- |  |
| CoronaVac × 3 | -- | 54 (42, 71) | -- |  |
| BBIBP-CorV × 2 + NVSI-06-08* |  | 368 (296, 457) | -- | Kaabi et al <sup>21</sup> |
| BBIBP-CorV × 3 | -- | 45 (36, 56) | -- |  |
| BBIBP-CorV × 2 + V-01*† | -- | 149 (102, 207) | -- | Wang et al <sup>20</sup> |
| CoronaVac × 2 + V-01*† | -- | 59 (54, 65) | -- |  |

\* Protein subunit vaccines
† Estimated from vaccine efficacy data (see Table S2)

**Table S2.**
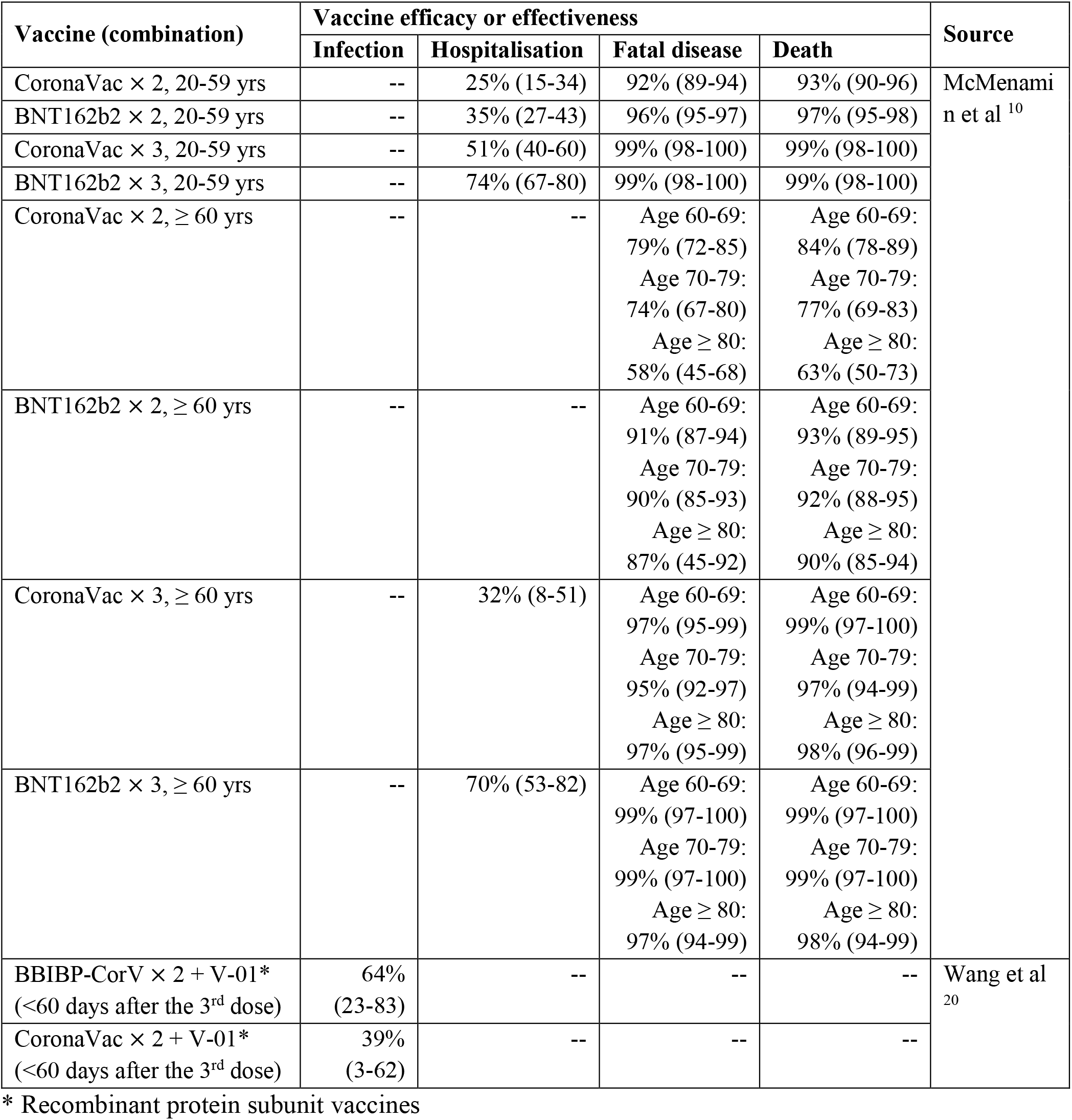
Vaccine efficacy or effectiveness in preventing Omicron infection, hospitalisation, fatal disease, and death.

**Table S3.** Model parameters.

| Parameter | Description, assumption, and source | Value |
| --- | --- | --- |
| $R_0$ | Basic reproductive number of Omicron BA.2 in the absence of vaccination <sup>16,33</sup> | BA.2: 7.1 (6.9-7.3)<br>BA.4/BA.5: 8.3 (7.8-8.9)<br>We estimate $R_0$ of BA.2 from the epidemiological data of the fifth wave in Hong Kong, and estimate $R_0$ of BA.4/BA.5 from the growth rates of England published by UKHSA as follows (https://www.gov.uk/guidance/the-r-value-and-growth-rate).<br>We assume that 86% of the England population had immunity against infection of BA.1, BA.2 or BA.2.12.1 by 1 May (i.e., $R_t = 1$ and $1-1/7.1 = 86\%$). In the week of 24 Jun, BA.4/BA.5 became the dominant variant and the daily growth rate was 2-5%, which corresponds to the relative increase in $R_t$ of 9.5-25.1%. The $R_0$ of BA.4/BA.5 is estimated to be 8.3 (7.8-8.9) accordingly. |
| $T_{GT}$ | Mean generation time <sup>34</sup> | 4.6 days |
| $f_{GT}$ | Probability density function of generation time <sup>34</sup> | Gamma (2.20, 2.09) |
| $VE_S$ | Vaccine effectiveness in reducing susceptibility | Table 1; Estimated by bootstrapping the neutralising antibody titres from Table S1 |
| $VE_I$ | Vaccine effectiveness in reducing infectiousness | Table 1; Assumed to be $0.8 \times VE_S$ |
| $VE_H$ | Vaccine effectiveness in reducing hospitalisation <sup>10</sup> | Table 1 |
| $VE_D$ | Vaccine effectiveness in reducing death <sup>10</sup> | Table 1 |
| $\varepsilon_S$ | Antiviral effectiveness in reducing susceptibility | 0 |
| $\varepsilon_I$ | Antiviral effectiveness in reducing infectivity | 0 |
| $\varepsilon_H$ | Antiviral effectiveness in reducing hospitalisations <sup>11</sup> | Nirmatrelvir/Ritonavir: 0.24 (0.14-0.33) |
| $\varepsilon_D$ | Antiviral effectiveness in reducing deaths <sup>11</sup> | Nirmatrelvir/Ritonavir: 0.66 (0.48-0.78) |
| $\sigma_{AR}$ | Coverage of antivirals among eligible patients <sup>16</sup> | 60%, estimated from preliminary data from Hong Kong |
| $p_{a,death}$ | Age-specific infection-fatality and infection-hospitalisation risk among unvaccinated individuals of a VOC similar to the Omicron variant <sup>35,36</sup> with no COVID-specific antivirals; assuming the hazard ratio of Delta variant was 1.45 times of that of Alpha variant and the hazard ratio of Omicron variant was 0.3 times of Delta variant <sup>16,37,38</sup> ; assuming Beta distributed with the coefficient of variation of 0.05. | Age 0-9: 0.0005%<br>Age 10-19: 0.0005%<br>Age 20-29: 0.0005%<br>Age 30-39: 0.023%<br>Age 40-49: 0.023%<br>Age 50-59: 0.126%<br>Age 60-69: 0.126%<br>Age 70-79: 2.00%<br>Age ≥ 80: 8.70% |
| $p_{a,hospitalisation}$ | | Age 0-9: 0.011%<br>Age 10-19: 0.027%<br>Age 20-29: 0.72%<br>Age 30-39: 2.34%<br>Age 40-49: 2.94%<br>Age 50-59: 5.52%<br>Age 60-69: 7.98%<br>Age 70-79: 11.28%<br>Age ≥ 80: 12.48% |
| $f_{incubation}$ | Probability density function of incubation period <sup>39,40</sup> | Lognormal distribution<br>Mean: 3.5 days<br>SD: 2.6 days |
| $f_{hospitalisation}$ | Probability density function of the time between infection and hospitalisation <sup>41</sup> | Gamma distribution<br>Mean: 6.7 days<br>SD: 3.0 days |
| $f_{death}$ | Probability density function of the time between infection and death; estimated from $f_{incubation}$ and the probability density function of the time between onset and death (Mean 18.8 days and SD 8.46 days) from Verity et al <sup>41</sup>; estimated from preliminary data from Hong Kong <sup>16</sup> | Gamma distribution<br>Mean: 22.3 days<br>SD: 9.5 days |
| $\tau_{hospitalisation}$ | The maximum daily incidence of hospitalisations that the health system could manage; assuming to be 100% and 200% of hospital beds designated for COVID patients in May 2022 in Hong Kong which correspond to its regular and surged capacity, respectively | 1.1 and 2.2 per 10,000 population<br>In late March 2022 when the daily incidence of hospitalisations peaked, the maximum number of hospital beds designated for COVID patients was 13,654 which corresponds to a maximum daily incidence of 2.28 hospitalisations per 10,000 population.<br>http://www.takungpao.com.hk/news/232109/2022/0325/701676.html<br>https://www.news.gov.hk/eng/2022/03/20220331/20220331_195929_222.html |
| $m_{death}$ | Increase in IFRs when health systems are overloaded | We assume that IFRs track the number of patients who require |
|  |  | hospital care: IFRs would increase by 10%, 20%, 30%, 40% and 50% when demand outstrips supply of available hospital beds by a ratio of 1-2, 2-3, 3-4, 4-5 and >5 to 1.<br>Our assumptions are based on data below:<br>(i) the estimated IFRs in Wuhan were about 50% higher than estimated IFRs in other provinces outside Hubei in 2020 <sup>42,43</sup>;<br>(ii) the incident rate ratios for hospital-based resources on COVID-19 deaths in the US between March and July 2020: geographical areas with fewer ICU beds (IRR = 0.194), nurses (IRR = 0.927) and general hospital beds (IRR = 0.800) per COVID-19 case were statistically associated with increased deaths in April 2020 <sup>44</sup>;<br>(iii) in-hospital mortality between March and August 2020 in the US: the adjusted mortality dropped from 25.6% in March 2020 to 7.6% in August 2020, and the standardized mortality ratio dropped from 1.26 in March to 0.38 in August 2020 <sup>45</sup>. |

**Table S4.** Effectiveness of PHSMs during the previous waves of COVID in Hong Kong and Shanghai.

| Second wave in Hong Kong (Ancestral strain Wuhan-Hu-1 and D614G, 29 February to 15 April 2020) |  |  |  |  |  |
| --- | --- | --- | --- | --- | --- |
| PHSM | Date | $R_t$ before | $R_t$ after | Reduction in $R_t$ | Level |
| School closure (entire 2<sup>nd</sup> wave) | 2/29 | 2.27<br>(1.70-2.93) | 1.28<br>(1.04-1.53) | 44%<br>(33-54) | 2 |
| Tightened travel control | 2/29-3/20 |  |  |  |  |
| Civil servants work-from-home | 3/21 | 1.28<br>(1.04-1.53) | 0.55<br>(0.38-0.70) | 76%<br>(69-83) | 3 |
| Tightened measures in restaurants and indoor amenities | 3/27 | 0.55<br>(0.38-0.70) | 0.20<br>(0.09-0.38) | 91%*<br>(83-96) | 4 |
| Closure of outdoor amenities | 3/27 |  |  |  |  |
| Closure of indoor amenities | 4/1 |  |  |  |  |
| Third wave in Hong Kong (Ancestral strain D614G, 30 June to 20 September 2020) |  |  |  |  |  |
| PHSM | Date | $R_t$ before | $R_t$ after | Reduction in $R_t$ | Level |
| Tightened measures in amenities | 7/9 | 2.60<br>(2.27-3.52) | 1.83<br>(1.54-2.23) | 30%<br>(14-41) | 2 |
| Closure of kindergartens and P1-3 | 7/10 |  |  |  |  |
| Gathering in groups of 4 only | 7/13 | 1.83<br>(1.54-2.23) | 1.55<br>(1.37-1.75) | 52%<br>(46-55) | 3 |
| Closure of all indoor amenities | 7/13 |  |  |  |  |
| No dine-in after 6 pm | 7/13 |  |  |  |  |
| Closure of all schools | 7/14 |  |  |  |  |
| Civil servants work-from-home | 7/20 | 1.55<br>(1.37-1.75) | 1.26<br>(1.16-1.40) |  |  |
| Gathering in groups of 2 only | 7/29 | 1.26<br>(1.16-1.40) | 0.57<br>(0.49-0.69) | 78%<br>(73-81) | 4 |
| No dine-in all day (lasted 2 days) | 7/29 |  |  |  |  |
| Fourth wave in Hong Kong (Ancestral strain D614G, 30 October 2020 to 31 January 2021) |  |  |  |  |  |
| PHSM | Date | $R_t$ before | $R_t$ after | Reduction in $R_t$ | Level |
| School closure (P1-P3, kindergarten) | 11/20 | 2.26<br>(1.99-2.79) | 1.76<br>(1.50-2.04) | 47%<br>(40-53) | 2 |
| Closure of singing and dancing venues incl. pubs and clubs | 11/20 |  |  |  |  |
| Closure of most indoor amenities | 11/24 | 1.76<br>(1.50-2.04) | 1.20<br>(1.06-1.36) |  |  |
| Closure of all schools | 11/29 | 1.20<br>(1.06-1.36) | 1.05<br>(0.97-1.16) | 55%<br>(51-59) | 3 |
| Civil servants work-from-home | 11/30 | 1.05<br>(0.97-1.16) | 1.01<br>(0.93-1.11) |  |  |
| No dine-in after 9 pm | 11/30 |  |  |  |  |
| No dine-in after 6 pm | 12/2 | 1.01<br>(0.93-1.11) | 0.69<br>(0.61-0.78) | 69%<br>(65-73) | 4 |
| Fifth wave in Hong Kong (Omicron BA.2, 1 January 2022 – Now) |  |  |  |  |  |
| PHSM | Date | $R_t$ before | $R_t$ after | Reduction in $R_t$ | Level |
| School closure | 1/5 | 7.1<br>(6.9-7.3) | 1.9<br>(1.8-2.0) | 73%<br>(71-75) | 4 |
| Closure of singing and dancing venues | 1/5 |  |  |  |  |
| Closure of most indoor amenities | 1/5 |  |  |  |  |
| Closure of all schools | 1/5 |  |  |  |  |
| Civil servants work-from-home | 1/5 |  |  |  |  |
| No dine-in after 6 pm | 1/5 |  |  |  |  |
| Banning gathering across families | 1/5 |  |  |  |  |
| Omicron BA.2 in Shanghai (1 March 2022 – Now) |  |  |  |  |  |
| PHSM | Date | $R_t$ before | $R_t$ after | Reduction in $R_t$ | Level |
| PHSMs with similar intensity of Level 4 in Hong Kong | 3/8 | 7.1<br>(6.9-7.3) | 2.4<br>(2.2-2.8) | 72%<br>(67-75) | 4 |
| Lockdown of communities with case reports (before city-wide lockdowns) | 3/11-3/26 |  | 2.0<br>(1.77-2.32) |  |  |

\* Overestimated because the outbreak died out

**Table S5.** Number of hospital beds per 10,000 in the 31 provinces in mainland China in 2020*.

| Province | Number of hospital beds | Number of hospital beds per 10,000 population | Maximum number of hospital beds designated to COVID-19 patients per 10,000† | Maximum daily incidence of hospitalisations that the health system could manage per 10,000† |
| --- | --- | --- | --- | --- |
| Beijing | 127033 | 58.0 | 8.7 | 1.1 |
| Tianjin | 68275 | 49.2 | 7.4 | 0.9 |
| Hebei | 441962 | 59.2 | 8.9 | 1.1 |
| Shanxi | 223650 | 64.1 | 9.6 | 1.2 |
| Inner Mongolia | 162072 | 67.4 | 10.1 | 1.3 |
| Liaoning | 314488 | 73.9 | 11.1 | 1.4 |
| Jilin | 173123 | 72.2 | 10.8 | 1.4 |
| Heilongjiang | 253345 | 79.9 | 12.0 | 1.5 |
| Shanghai | 152191 | 61.2 | 9.2 | 1.2 |
| Jiangsu | 535006 | 63.1 | 9.5 | 1.2 |
| Zhejiang | 361317 | 55.9 | 8.4 | 1.1 |
| Anhui | 407813 | 66.8 | 10.0 | 1.3 |
| Fujian | 216753 | 52.1 | 7.8 | 1.0 |
| Jiangxi | 285847 | 63.3 | 9.5 | 1.2 |
| Shandong | 646863 | 63.6 | 9.5 | 1.2 |
| Henan | 667156 | 67.1 | 10.1 | 1.3 |
| Hubei | 411351 | 71.6 | 10.7 | 1.3 |
| Hunan | 519902 | 78.2 | 11.7 | 1.5 |
| Guangdong | 564773 | 44.7 | 6.7 | 0.8 |
| Guangxi | 295562 | 58.9 | 8.8 | 1.1 |
| Hainan | 58474 | 57.8 | 8.7 | 1.1 |
| Chongqing | 235520 | 73.4 | 11.0 | 1.4 |
| Sichuan | 649756 | 77.6 | 11.6 | 1.5 |
| Guizhou | 276379 | 71.6 | 10.7 | 1.3 |
| Yunnan | 325212 | 68.9 | 10.3 | 1.3 |
| Tibet | 18586 | 50.8 | 7.6 | 1.0 |
| Shaanxi | 272424 | 68.9 | 10.3 | 1.3 |
| Gansu | 171866 | 68.7 | 10.3 | 1.3 |
| Qinghai | 41285 | 69.6 | 10.4 | 1.3 |
| Ningxia | 41261 | 57.2 | 8.6 | 1.1 |
| Xinjiang | 181455 | 70.1 | 10.5 | 1.3 |

\* Data from https://data.cnki.net/Trade/Home/Index/Z020
† Assumed to be 15% of total number of hospital beds and the average length of stay is 8 days

**Table S6.** Number of secondary and tertiary hospitals in the 31 provinces in mainland China in 2020*.

| Province | Number of secondary hospitals | Number of tertiary hospitals | Number of secondary hospitals per 1,000,000 | Number of tertiary hospitals per 1,000,000 |
| --- | --- | --- | --- | --- |
| Beijing | 158 | 106 | 7.2 | 4.8 |
| Tianjin | 76 | 43 | 5.5 | 3.1 |
| Hebei | 601 | 99 | 8.1 | 1.3 |
| Shanxi | 381 | 61 | 10.9 | 1.7 |
| Inner Mongolia | 314 | 87 | 13.1 | 3.6 |
| Liaoning | 338 | 156 | 7.9 | 3.7 |
| Jilin | 280 | 53 | 11.6 | 2.2 |
| Heilongjiang | 364 | 102 | 11.4 | 3.2 |
| Shanghai | 104 | 44 | 4.2 | 1.8 |
| Jiangsu | 470 | 192 | 5.5 | 2.3 |
| Zhejiang | 220 | 137 | 3.4 | 2.1 |
| Anhui | 513 | 98 | 8.4 | 1.6 |
| Fujian | 272 | 87 | 6.5 | 2.1 |
| Jiangxi | 256 | 88 | 5.7 | 1.9 |
| Shandong | 714 | 197 | 7.0 | 1.9 |
| Henan | 605 | 115 | 6.1 | 1.2 |
| Hubei | 361 | 143 | 6.3 | 2.5 |
| Hunan | 519 | 107 | 7.8 | 1.6 |
| Guangdong | 560 | 231 | 4.4 | 1.8 |
| Guangxi | 311 | 85 | 6.2 | 1.7 |
| Hainan | 58 | 36 | 5.8 | 3.6 |
| Chongqing | 263 | 61 | 8.2 | 1.9 |
| Sichuan | 741 | 269 | 8.9 | 3.2 |
| Guizhou | 364 | 65 | 9.4 | 1.7 |
| Yunnan | 470 | 106 | 10.0 | 2.2 |
| Tibet | 53 | 14 | 14.5 | 3.8 |
| Shaanxi | 420 | 73 | 10.6 | 1.8 |
| Gansu | 210 | 47 | 8.4 | 1.9 |
| Qinghai | 88 | 24 | 14.9 | 4.1 |
| Ningxia | 87 | 15 | 12.1 | 2.1 |
| Xinjiang | 233 | 55 | 9.0 | 2.1 |

\* Data from https://data.cnki.net/Trade/Home/Index/Z020

**Table S7.** Proportion of sub-administrative regions coded as “rural” in cities and counties in Henan, Guangxi, and Guangdong (PRD).

| City/county | Code of city/county | Number of sub-administrative regions | Number of urban sub-administrative regions | Number of rural sub-administrative regions | Proportion of rural sub-administrative regions |
| --- | --- | --- | --- | --- | --- |
| **Henan** |  |  |  |  |  |
| Zhengzhou* | 4101 | 3206 | 1739 | 1467 | 46% |
| Kaifeng | 4102 | 2573 | 916 | 1657 | 64% |
| Luoyang | 4103 | 3228 | 984 | 2244 | 70% |
| Pingdingshan | 4104 | 2828 | 759 | 2069 | 73% |
| Anyang | 4105 | 3300 | 933 | 2367 | 72% |
| Hebi | 4106 | 1030 | 413 | 617 | 60% |
| Xinxiang | 4107 | 3820 | 1219 | 2601 | 68% |
| Jiaozuo | 4108 | 2021 | 807 | 1214 | 60% |
| Puyang | 4109 | 3201 | 895 | 2306 | 72% |
| Xuchang | 4110 | 2492 | 816 | 1676 | 67% |
| Luohe | 4111 | 1331 | 423 | 908 | 68% |
| Sanmenxia | 4112 | 1379 | 324 | 1055 | 77% |
| Nanyang | 4113 | 4960 | 1270 | 3690 | 74% |
| Shangqiu | 4114 | 4834 | 1269 | 3565 | 74% |
| Xinyang | 4115 | 3463 | 775 | 2688 | 78% |
| Zhoukou | 4116 | 5099 | 1452 | 3647 | 72% |
| Zhumadian | 4117 | 2906 | 550 | 2356 | 81% |
| **Guangxi** |  |  |  |  |  |
| Nanning* | 4501 | 1845 | 614 | 1231 | 67% |
| Liuzhou | 4502 | 1237 | 461 | 776 | 63% |
| Guilin | 4503 | 1924 | 444 | 1480 | 77% |
| Wuzhou | 4504 | 1015 | 318 | 697 | 69% |
| Beihai | 4505 | 436 | 122 | 314 | 72% |
| Fangchenggang | 4506 | 349 | 84 | 265 | 76% |
| Qinzhou | 4507 | 1042 | 213 | 829 | 80% |
| Guigang | 4508 | 1179 | 349 | 830 | 70% |
| Yulin | 4509 | 1510 | 434 | 1076 | 71% |
| Baise | 4510 | 1897 | 265 | 1632 | 86% |
| Hezhou | 4511 | 761 | 225 | 536 | 70% |
| Hechi | 4512 | 1661 | 231 | 1430 | 86% |
| Laibin | 4513 | 822 | 186 | 636 | 77% |
| Chongzuo | 4514 | 932 | 147 | 785 | 84% |
| **Guangdong** |  |  |  |  |  |
| Guangzhou* | 4401 | 2827 | 2136 | 691 | 24% |
| Shaoguan | 4402 | 1476 | 459 | 1017 | 69% |
| Shenzhen | 4403 | 784 | 784 | 0 | 0% |
| Zhuhai | 4404 | 339 | 254 | 85 | 25% |
| Shantou | 4405 | 1089 | 666 | 423 | 39% |
| Foshan | 4406 | 815 | 736 | 79 | 10% |
| Jiangmen | 4407 | 1334 | 522 | 812 | 61% |
| Zhanjiang | 4408 | 1986 | 565 | 1421 | 72% |
| Maoming | 4409 | 1916 | 747 | 1169 | 61% |
| Zhaoqing | 4412 | 1553 | 415 | 1138 | 73% |
| Huizhou | 4413 | 1345 | 487 | 858 | 64% |
| Meizhou | 4414 | 2267 | 696 | 1571 | 69% |
| Shanwei | 4415 | 907 | 349 | 558 | 62% |
| Heyuan | 4416 | 1443 | 336 | 1107 | 77% |
| Yangjiang | 4417 | 871 | 289 | 582 | 67% |
| Qingyuan | 4418 | 1230 | 331 | 899 | 73% |
| Dongguan | 4419 | 608 | 572 | 36 | 7% |
| Zhongshan | 4420 | 279 | 235 | 44 | 16% |
| Chaozhou | 4451 | 1035 | 421 | 614 | 59% |
| Jieyang | 4452 | 1651 | 647 | 1004 | 61% |
| Yunfu | 4453 | 980 | 339 | 641 | 65% |

\* Provincial capital

**Figure S1.**
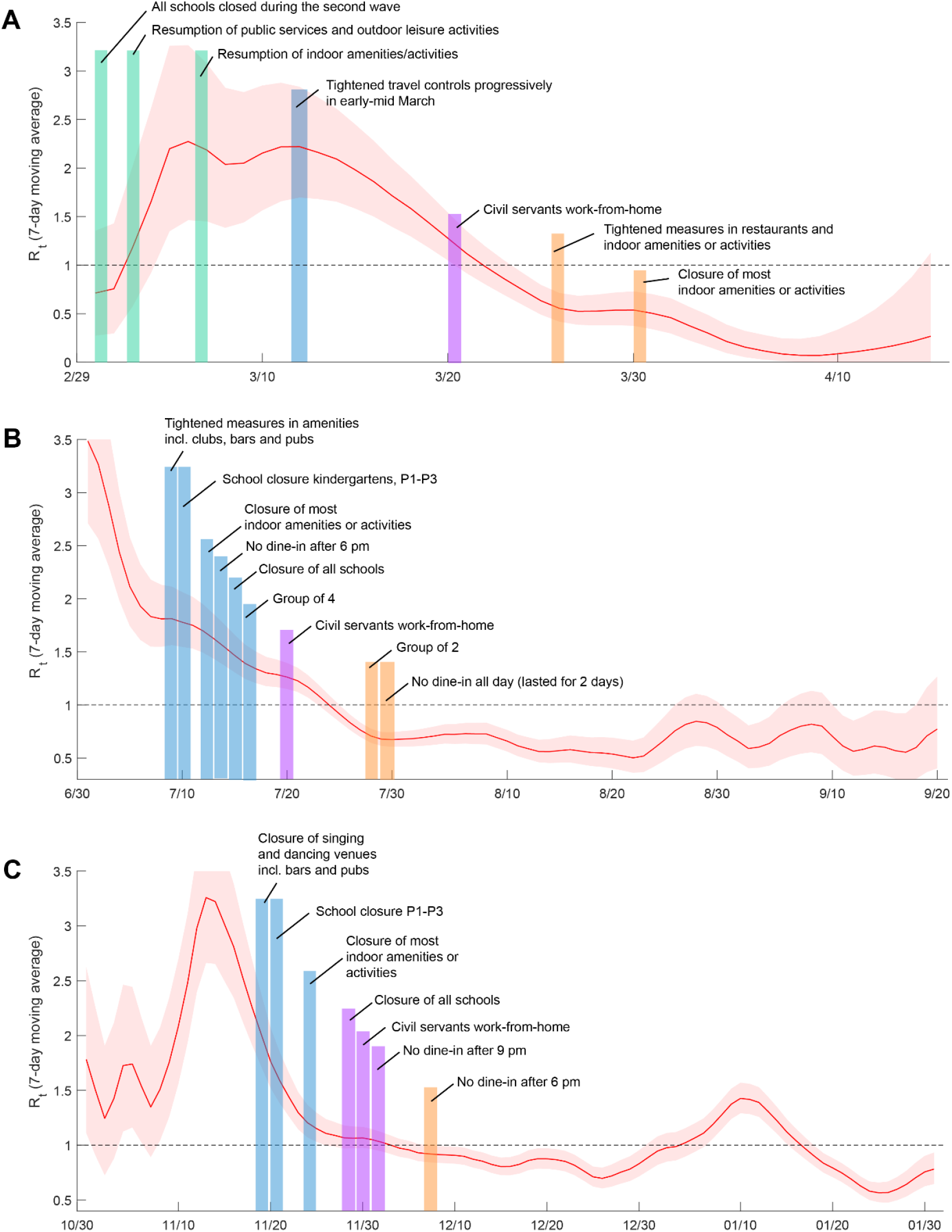
Correlation between *R*_*t*_ and public health and social measures (PHSMs) implemented in the second, third and fourth wave in Hong Kong. *R*_*t*_ were estimated from deconvoluted time series of daily number of cases in the EpiEstim model ^43^.

**Figure S2.**
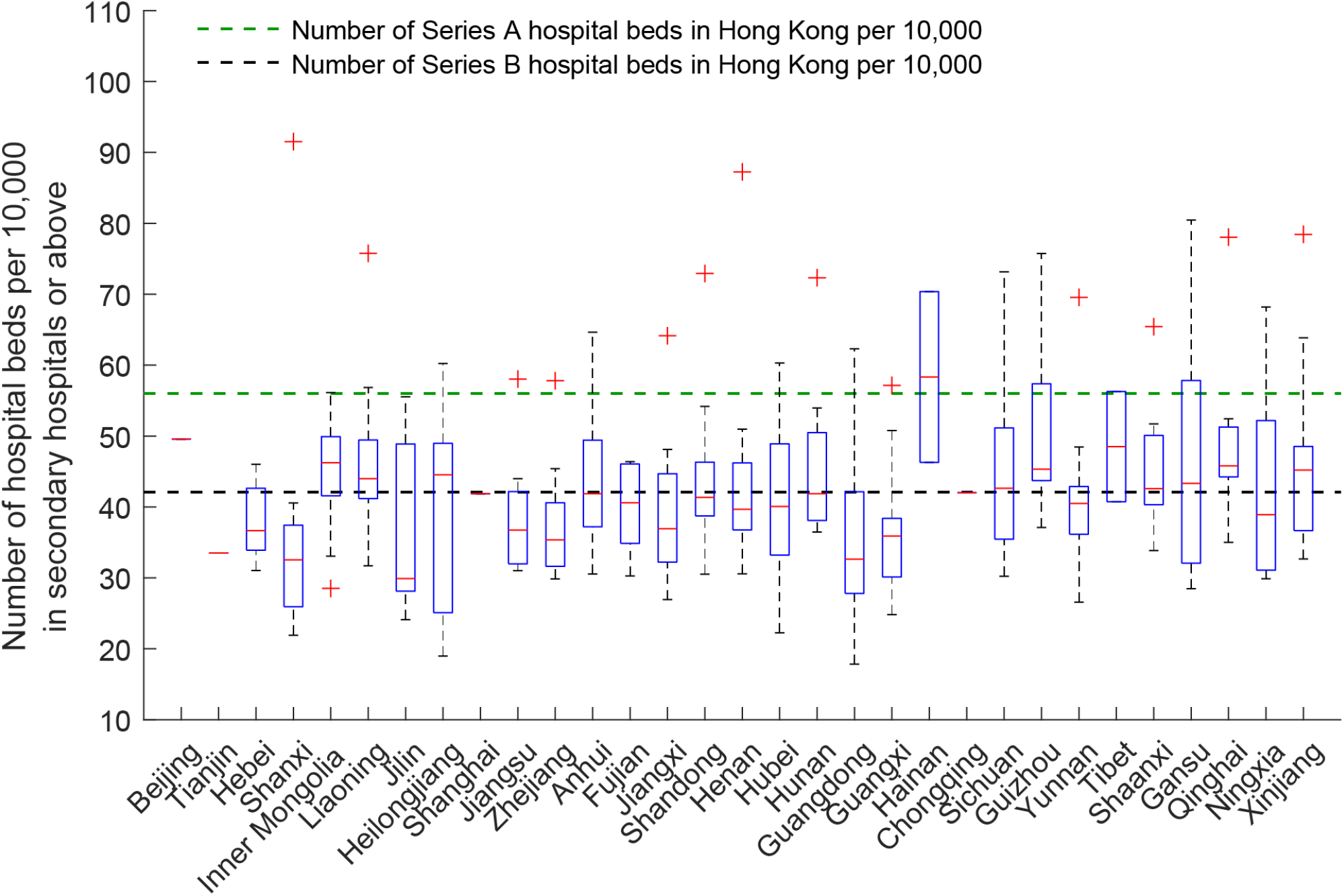
Number of hospital beds per 10,000 in secondary hospitals or above in more than 300 cities in mainland China. Cities are grouped by provinces where they are located. Hospitals in mainland China are classified into three levels including primary, secondary, and tertiary hospitals: 1) A primary hospital is typically a township hospital that contains less than 100 beds, and primary hospitals provide preventive care, minimal health care and rehabilitation services; 2) A secondary hospital is one that tend to be affiliated with a medium size city, county or district and contain more than 100 beds but less than 500 and secondary hospitals are responsible for providing comprehensive health services; 3) A tertiary hospital is a comprehensive, referral, general hospitals at the city, provincial or national level with a bed capacity exceeding 500 and tertiary hospitals serve as medical hubs providing care to multiple regions. The national average number of hospital beds in secondary hospitals or above is 42 per 10,000. The two dashed lines show the number of hospital beds of Service A (56 per 10,000) and Service B (42 per 10,000) in 2020 in Hong Kong (https://www.fhb.gov.hk/statistics/en/health_statistics.htm). Series A included all hospital beds in Hospital Authority hospitals, private hospitals, nursing homes and correctional institutions. Series B included only hospital beds in Hospital Authority hospitals and private hospitals excluding accident and emergency observation beds, day beds and nursery beds, which followed the definition of OECD Health Data.

**Figure S3.**
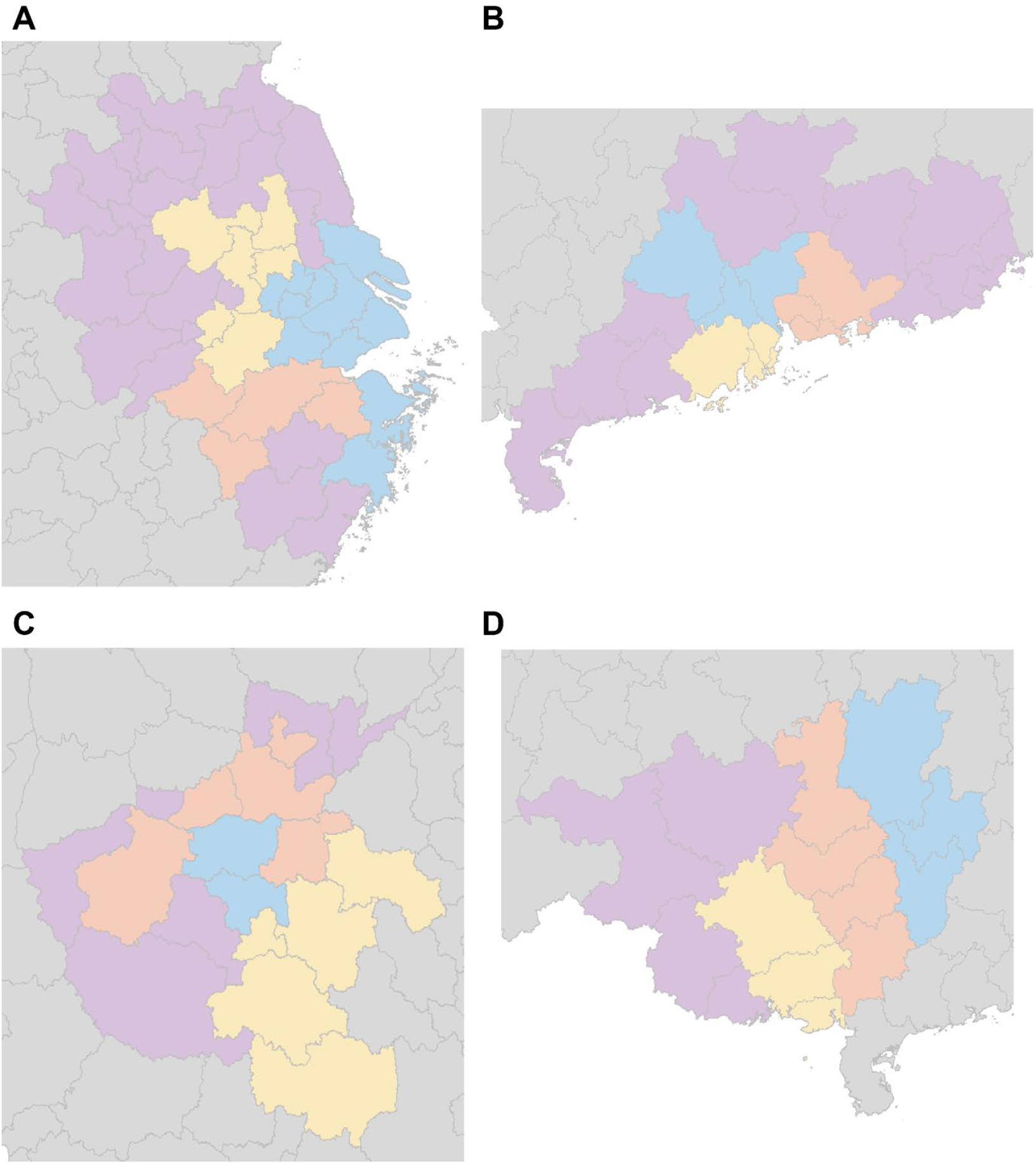
Simultaneous reopening of Yangtze River Delta Region, Pearl River Delta Region, Henan, and Guangxi. (A) Yangtze River Delta: Yangtze River Delta is assumed to be reopened simultaneously in Greater Shanghai (blue), Greater Hangzhou (orange), Greater Nanjing (yellow) and all cities in Anhui, Jiangsu, and Zhejiang (purple). **(B) Pearl River Delta:** Pearl River Delta is assumed to be reopened simultaneously in Guangzhou-Foshan-Zhaoqing (blue), Shenzhen-Dongguan-Huizhou (orange), Zhuhai-Zhongshan-Jiangmen (yellow) and all cities in Guangdong (purple). **(C) Henan:** Henan is assumed to be reopened simultaneously in provincial capital area (blue), urban area (orange), urban-rural junction (yellow) and rural area (purple). **(D) Guangxi:** Guangxi is assumed to be reopened simultaneously in the areas adjacent to the most developed parts of Guangdong (blue), the central areas of Guangxi (orange), the provincial capital area (yellow), and the areas next to China-Vietnam border (purple).

**Figure S4.**
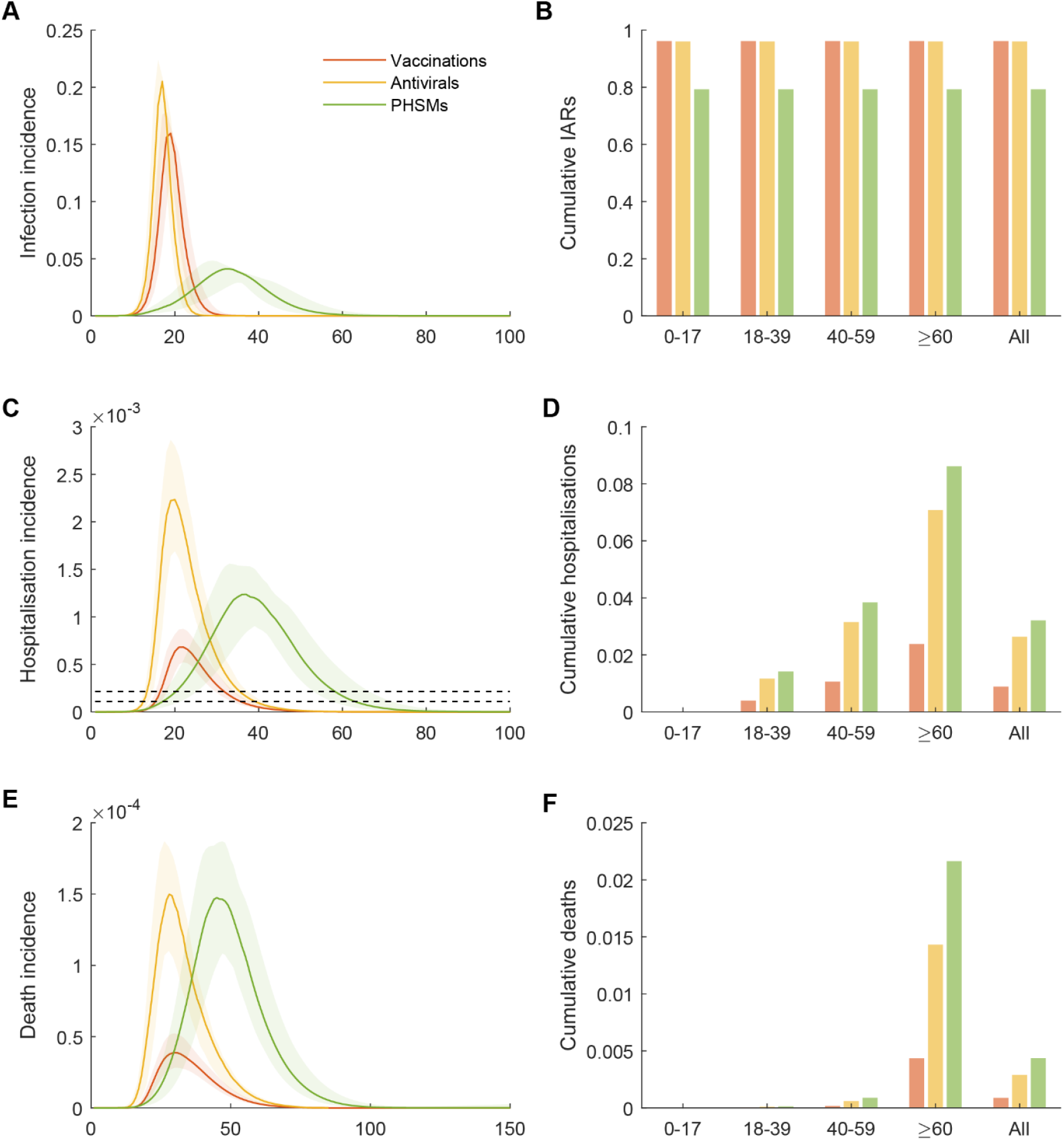
The impacts of 4th dose heterologous boosting, antiviral treatments and PHSMs if they are singly implemented during reopening under Strategy 1. We assume that the three interventions would be implemented follows: 1) Increasing the vaccine uptake of the 3rd and 4th dose to 85% across all age groups aged 3 or above and the mass vaccination of the 4th dose starts 30 days before the reopening; 2) Increasing the antiviral coverage to 60%; 3) PHSMs at Level 4 which reduce *R*_*t*_ by 69-72% are implemented 14 days after the seeding of epidemics, and PHSMs are maintained between 15 and 74 days after the seeding of epidemics, and gradually relaxed between 75 and 104 days. **(A) Infection incidence as proportion of the population. (B) Cumulative infection attack rates by age. (C-D) Daily and cumulative number of cases requiring who require hospitalisation. (E-F) Daily and cumulative incidence of death**. Figure legend and other parameters are the same as Figure 2.

## Notes

### Competing Interest Statement

The authors have declared no competing interest.

